# Alpha-synuclein misfolding as a fluid biomarker for Parkinson’s disease and synucleinopathies measured with the iRS platform

**DOI:** 10.1101/2024.09.02.24312694

**Authors:** Martin Schuler, Grischa Gerwert, Marvin Mann, Nathalie Woitzik, Lennart Langenhoff, Diana Hubert, Deniz Duman, Adrian Höveler, Sandy Galkowski, Jonas Simon, Robin Denz, Sandrina Weber, Eun-Hae Kwon, Robin Wanka, Carsten Kötting, Jörn Güldenhaupt, Léon Beyer, Lars Tönges, Brit Mollenhauer, Klaus Gerwert

**Affiliations:** Center for Protein Diagnostics (PRODI), Ruhr-University Bochum, Bochum, Germany; Department of Biophysics, Ruhr-University Bochum, Bochum, Germany; Department of Medical Informatics, Biometry and Epidemiology, Ruhr-University Bochum, Germany; University Medical Center Göttingen, Department of Neurology, Göttingen, Germany; Department of Neurology, St. Josef-Hospital, Ruhr-University Bochum, Bochum, Germany; Paracelsus-Elena-Klinik, Kassel, Germany (Klinikstraße 16, 34128 Kassel, Germany); Scientific employee with an honorary contract at Deutsches Zentrum für Neurodegenerative Erkrankungen (DZNE), Göttingen, Germany

## Abstract

Misfolding and aggregation of alpha-synuclein (αSyn) plays a key role in the pathophysiology of Parkinson’s disease (PD). It induces cellular and axonal damage already in the early stages of the disease. Despite considerable advances in PD diagnostics by αSyn seed-amplification assays (SAAs), an early and differential diagnosis of PD still represents a major challenge. Here, we extended the immuno-infrared sensor (iRS) platform technology from Alzheimer’s disease (AD), in which β-amyloid misfolding was monitored as a fluid biomarker towards αSyn misfolding in PD. Using the iRS platform technology, we analyzed cerebrospinal fluid (CSF) from two independent cohorts, a discovery and a validation cohort comprising clinically diagnosed PD (n=57), atypical Parkinsonian disorders with αSyn pathology (multiple system atrophy (MSA), n= 5) or Tau pathology (corticobasal degeneration (CBD), n=5, progressive supranuclear palsy (PSP) n=9), and further disease controls (frontotemporal dementia (FTD) n=7 and other, n=51). In the discovery cohort, an AUC of 0.90, 95 %-CL 0.85 – 0.96 is obtained for the differentiation of PD/MSA vs. all controls, and in the validation cohort, an AUC of 0.86, 95 %-CL 0.80 - 0.93, respectively. In the combined dataset, the αSyn misfolding classifies PD/MSA from controls with an AUC of 0.90 (n=134, 95 %-CL 0.85 - 0.96). Using two threshold values instead of one identified people in the continuum between clearly unaffected (low misfolding group) and affected by PD/MSA (high misfolding group) with an intermediate area in between. The controls versus PD/MSA in the low vs. high misfolding group were classified with 97% sensitivity and 92% specificity.

The spectral data showed misfolding in CSF from an α-helical/random-coil secondary structure of αSyn in controls to β-sheet enriched secondary structures in PD and MSA patients. In subgroups, the iRS platform implied a potential for stratifying patients with overlapping clinical symptoms. With high accuracy, the iRS αSyn misfolding platform provides a novel diagnostic tool using body fluids for differential, biological classification of the αSyn associated disorders PD and MSA. The iRS platform, indicating directly and fast all αSyn conformers, is complementary to the αSyn SAAs, which also uses αSyn misfolding as a fluid biomarker. However, monomers amplify competent misfolded conformers in SAA over time. The iRS platform opens new avenues for the stratification of PD by a body-fluid analysis and follow-ups in the continuum of healthy to clinically impaired individuals.

## Introduction

Parkinson’s disease (PD) is a frequent neurodegenerative disorder that causes significant disability and an increasing global public health burden related to motor and non-motor features.^1^ It is characterized by a progressive loss of dopaminergic and non-dopaminergic neurons in the CNS.^2,3^

Current PD diagnostic criteria mainly rely on the onset of clinical hallmark motor symptoms, including bradykinesia, rigidity, resting tremor, and postural instability. The disease is often advanced at the time of symptom onset, and over 50 % of nigral dopaminergic neurons are already lost.^4^ Unlike Alzheimer’s disease (AD), blood-based biomarkers and positron emission tomography (PET) for diagnosis are subject to ongoing research and have not yet been well established for αSyn.^5,6^ Therefore, objective biomarkers that accurately verify PD and distinguish it from other Parkinsonian disorders, ideally in the prodromal or earliest disease stages, are urgently needed and under discussion.^7,8^

The neuropathological hallmark of PD is the abnormal accumulation of αSyn in Lewy bodies.^9^ Various factors, such as genetic predisposition and post-translational modifications, are believed to contribute to the misfolding and aggregation of αSyn, resulting in the formation of oligomers, amyloid-like fibrils, and deposits such as Lewy bodies.^10^ In recent years, new body fluids techniques, including protein amplification assays such as Protein Misfolding Cyclic Amplification (PMCA) and the Real-Time Quaking-Induce Conversion (RTQuIC), now known under the consensus term of seed amplification assays (SAA), have emerged.^11–13^ Previous studies have shown that these techniques detect aggregated and misfolded αSyn in cerebrospinal fluid (CSF) in clinical stages with accuracies over 90 %.^14,15^ However, these assays strongly rely on multiple factors (matrix, wells, buffers, etc.), including the quality of the reaction substrate for amplification, and comparability for quantification needs further harmonization, which is ongoing.^16,17^

Complementary, the immuno-infrared-sensor (iRS) platform measures the misfolding of the biomarker directly and label-free by use of infrared spectroscopy. This provides a direct measure of the secondary structure distribution of the respective fluid biomarker by difference spectroscopy. Suppose the β-sheet misfolded conformers of β-amyloid in AD, are predominant compared to the α-helical/random-coil conformers. In that case, it indicates a high risk for a later AD clinical diagnosis in an early symptom-free stage.^18^ Misfolding of β-amyloid as a fluid biomarker for high AD risk is shown in individuals with AD, MCI, subjective cognitive decline, and for symptom-free stages.^19–23^ Importantly, past iRS studies for β-amyloid demonstrated robust CSF and blood matrix performance. In a real-world community-based cohort, high risk for clinical AD diagnosis was determined up to 17 years in advance in a symptom-free stage.^24^

The present study extends the iRS approach to αSyn misfolding in CSF in discovery and an independent validation study for identifying individuals with PD, atypical Parkinsonian syndromes, and disease controls.

## Methods

### Antigen preparations

Generation of αSyn pre-formed-fibrils (PFFs) was conducted according to the protocol of Polinski et al.^25^ Briefly, αSyn stocks (Stressmarq, Cat. No. SPR-321), stored at -80 °C, were slowly thawed on ice. The protein solution was centrifuged at 14400 x*g* for ten minutes at 4 °C to remove aggregates. The protein concentration of the supernatant was determined using the Bradford assay. Monomeric reference material aliquots were prepared in 1x Dulbecco’s PBS (Invitrogen 14190136) with a concentration of 6.9 μM or concentrated for ThT-measurement (346 µM), snap frozen in liquid N_2,_ and stored at -80 °C. Aliquots were only used once and diluted in PBS (pH 7.4) immediately before use and to the desired concentration.

For the generation of PFFs, peptide films were prepared to 346 μM in PBS, mixed, and incubated for 7 days at 37 °C under constant shaking at 1000 rpm. Successful fibrilization was reviewed by thioflavin T assay (**compare ESI Figure 1S**) and by secondary-structure sensitive infrared spectroscopy against the buffer background. PFFs aliquots were snap-frozen in liquid N_2_ and long-term stored at -80°C or ambient temperature for use within one week. Next to the inhouse-produced fibrils, PFFs type I (Cat. No. SPR-322) from Stressmarq were acquired as reference material for comparison. In addition, human recombinant αSyn-oligomers (dopamine-HCl stabilized; Stressmarq. Cat. No. SPR-466) were aliquoted and stored at -80°C upon arrival.

**Figure 1.**
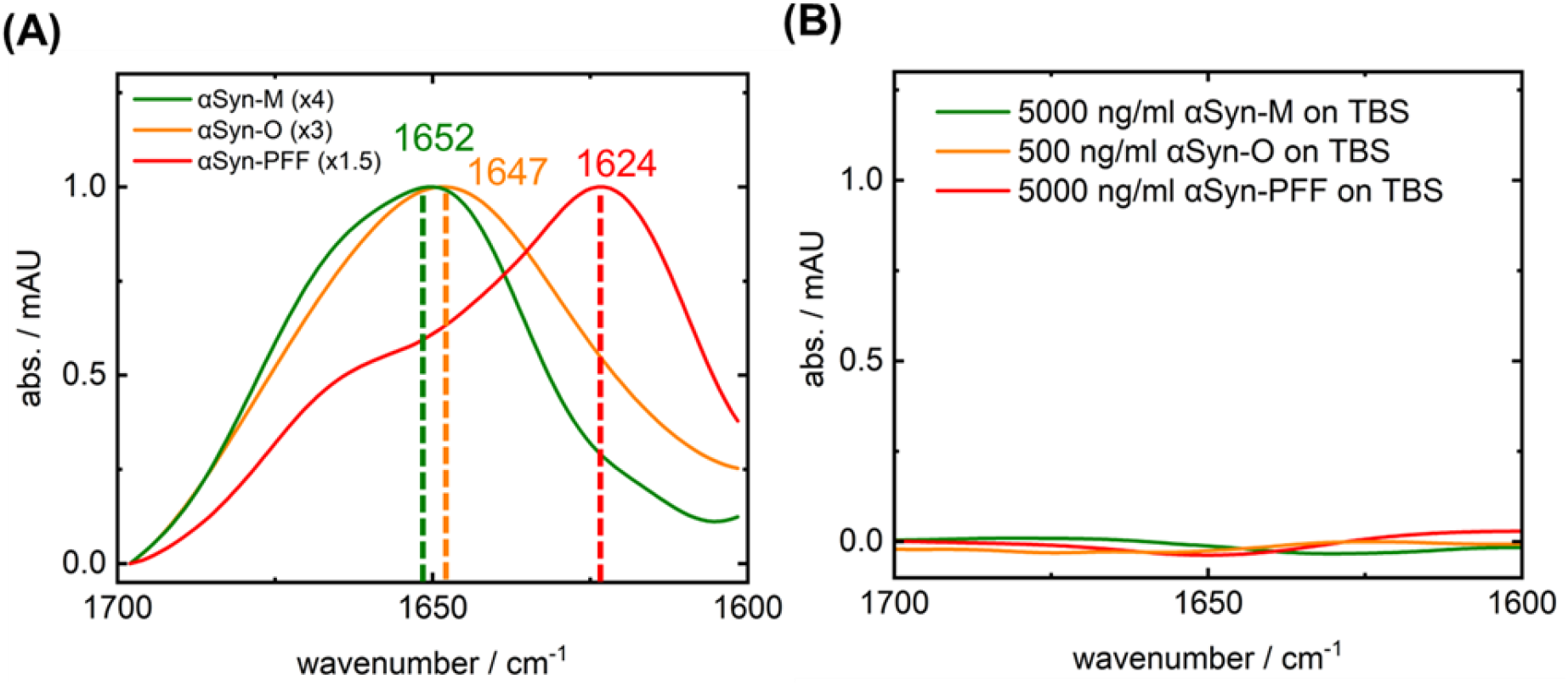
iRS-surface characterization by synthetic αSyn antigens. **Panel A**: The secondary structure sensitive Amide-I band absorbance of capture-antibody-bound αSyn-monomers (green, Stressmarq Bioscience Inc SPR-321), αSyn-oligomers (orange, Stressmarq Bioscience Inc SPR-466), and αSyn-PPFs (red, Stressmarq Bioscience Inc SPR-322) in PBS at high concentration of 500 ng/ml and scaled (x1.5-4) for comparison. Their structural differences are indicated by the significant wavenumber shift (cm^-1^) ranging from 1652 cm^-1^ for α-helical/random-coil (green) over 1647 cm^-1^ (orange) to β-sheet dominated structures absorbing at 1624 cm^-1^ (red). **Panel B**: The inertness measures on the blocking solution (BS) layer at high concentrations (500-5000 ng/ml) without the capture antibody. No signal is observed without the antibody on the blocking layer demonstrating sufficient inertness for pg-ng/ml concentrations of αSyn in CSF. Abbreviations: αSyn, alpha-synuclein; BS, blocking solution; PFF, pre-formed fibril.

### Study cohorts

CSF samples of the discovery study were collected following standard diagnostic, clinical GCP, and current research guidelines and processed as previously described.^26–28^ CSF samples of the validation study were collected from one German PD specialty center using SOPs as published before (Paracelsus-Elena-Klinik Kassel, Germany).^29^ Samples did not have artificial blood admixtures. The clinical diagnoses of Parkinson’s disease (PD), multiple system atrophy (MSA), progressive supranuclear palsy (PSP), corticobasal degeneration (CBD), and frontotemporal dementia (FTD) were made according to current consensus criteria.^30–35^ All samples are listed in detail in **ESI Table 1S**.

**Table 1.** Study cohorts and sample demographics.

| Parameter | PD/MSA |  |  | control |  |  | Statistics PD/MSA vs. control |  |  |
| --- | --- | --- | --- | --- | --- | --- | --- | --- | --- |
|  | Dis. | Val. | Comb. | Dis. | Val. | Comb. | Dis. | Val. | Comb. |
| **N total** | 17<br>17=PD | 45<br>40=PD<br>5=MSA | 62 | 42=control | 30<br>5=CBD<br>7=FTD<br>9=PSP<br>9=control | 72 | - | - | - |
| **Age**<br>[median±SD] | 69 | 70 | 70 | 64 (*) | 75 | 70 | MWU<br>(p = .243) | MWU<br>(p = .181) | MWU<br>(p = .540) |
| **Ratio of female [%]** | 18 | 25 | 23 | 58 | 48 | 53 | Chi<sup>2</sup><br>p <.0001 | Chi<sup>2</sup><br>p .0007 | Chi<sup>2</sup><br>p .0001 |
| **H&Y Scale %**<br>[mean±SD] | 100<br>[2.6±0.9] | 35<br>[3.2±0.9] | 52<br>[2.9±0.9] | 5<br>[3.5±0.5] | 11<br>[2.3±0.5] | 7<br>[2.8±0.75] | - | - | - |
| **UPDRS-III %**<br>[mean±SD] | 23<br>[26±10] | 94<br>[32±10] | 74<br>[32±10] | - | 48<br>[41±14] | 19<br>[41±14] | - | - | - |
| **Q<sub>Alb</sub> pos.-rate %**<br>[mean±SD]<br>(**) | 53<br>[9.5±5.3]<br>data<br>17/17 | 57<br>[10.4±5.6]<br>data<br>42/48 | 56<br>[10.1±5.5]<br>data<br>59/65 | 27<br>[6.7±3.0]<br>data<br>15/42 | 19<br>[7.7±3.9]<br>data<br>19/27 | 21<br>[7.3±3.5]<br>data<br>34/69 | - | - | - |

Abbreviations: BBB, blood-brain barrier; CBD, corticobasal degeneration; control, disease control; FTD, frontotemporal dementia; H&Y, Hoehn & Yahr; MSA, multiple system atrophy; MWU, Mann-Whitney-U; N, number of participants; PD, Parkinson's disease; UPDRS, Unified Parkinson's Disease Rating Scale, (\*) Nine cases w/o age/sex data, (\*\*) Age-dependent albumin CSF/Serum quotient ( $Q_{Alb} = Alb_{CSF}/Alb_{Serum}$ ) for assessment of BBB-dysfunction. Positivity for $Q_{Alb}$ was assumed when CSF/serum albumin quotient was higher than the calculated norm-value according to equation $Alb_{norm} = (4 + (age/15))$ .<sup>36,37</sup>

Study subjects gave their informed consent, and study approval was obtained by the local ethics committee and institutional review boards (Bochum cohort institutional review board (IRB) number: # 17-6119; Kassel cohort: IRB Vote from Landesärztekammer Hessen: FF89/2008 and FF38/2016). Further samples have been obtained from routine outpatient care in a fully anonymized procedure for which the responsible ethics committee (Ethik-Kommission der Ärztekammer Westfalen-Lippe und der Westfälischen Wilhelms-Universität Münster, Germany) has waived ethical approval (file number 2022-692-f-N).

**Table 1** summarizes the study cohorts and demographics, including age, sex, diagnosis, and additional information (H&Y scale, UPDRS-III, Albumin-Ratio (Q_Alb_)) upon availability. CSF samples were collected in the morning under fasting conditions, centrifuged, aliquoted, and stored at -80 °C on clinical sides. For iRS analysis, single-use aliquots (300 µl) of all CSF samples were provided and measured in a blinded manner. After data acquisition and analysis, results were reported to cooperation sites, and clinical diagnosis and additional information were provided for performance evaluation.

### Determination of αSyn-misfolding by the iRS platform

In this approach, the structure-sensitive amide-I band of αSyn is read out in the infrared spectral region. This band reflects the C=O stretching vibration of the αSyn peptide backbone. The frequency of this band is downshifted when αSyn misfolds because the structure changes from a mainly random-coil/α-helical to β-sheet enriched secondary structure.^38,39^ To subtract the large water background absorbance masking the much smaller αSyn absorbance, the attenuated total reflection (ATR) technology is applied. In the ATR approach, the evanescent wave generated by incident infrared light invades only about 500 nm of body fluid. This thin layer absorbance allows the subtraction of the water background absorbance, which is comparably large to sample signals and reveals by difference spectroscopy the αSyn absorbance spectrum (**compare ESI Figure 2S**).

**Figure 2.**
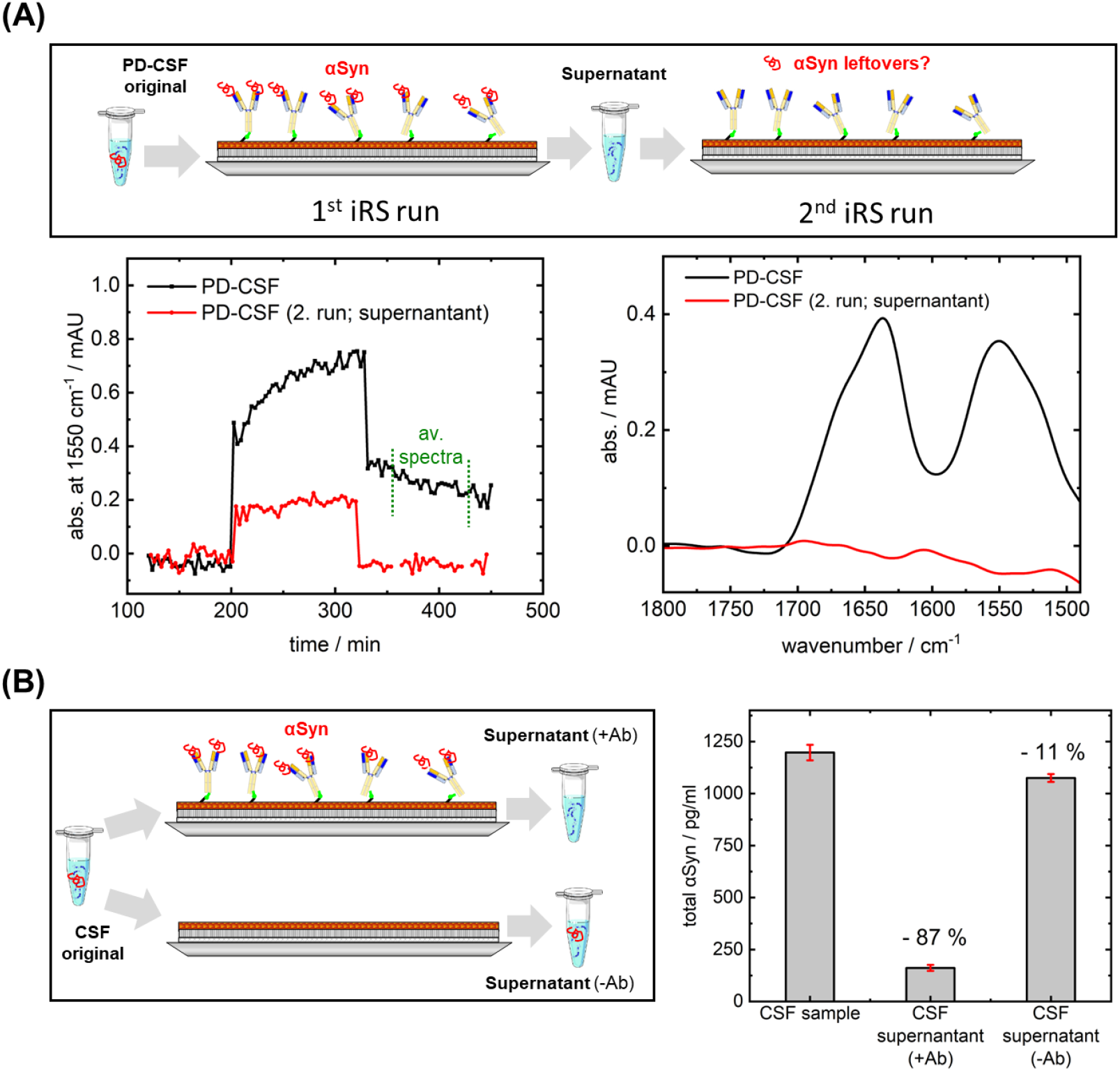
iRS CSF sample kinetics and spectra of two subsequent runs of the same sample and indirect-ELISA experiments for quantification of αSyn reduction in samples with iRS-setup. **Panel A**: CSF sample signal as kinetic and averaged results spectra (averaging minutes 10-110 of wash) in two consecutive runs. The kinetic and sample signals of the first run show a signal, while the second run, regardless of the predilution, does not show any binding in the sample wash. **Panel B**: The setup of the ELISA experiment, where a CSF sample was circulated on a surface with and without the capture antibody. The original sample was withheld to be compared to the CSF supernatant collected after sample circulation. Dilution factors of iRS analysis were considered where needed. The original sample contained 1197 ± 37 pg/ml αSyn, while the supernatant of the CSF circulated in the presence of the capture antibody marks a decreased αSyn concentration of 161 ± 14 pg/ml. The αSyn concentration in supernatant without the capture antibody on the blocked surface is slightly reduced to 1075 ± 19 pg/ml. Samples and supernatants were diluted according to the standard range and measured in duplicates. The standard deviation of duplicates is reported as error bars (red). Abbreviations: Ab, antibody; αSyn, alpha-synuclein; CSF, cerebrospinal fluid; iRS, immuno-infrared-sensor.

The ATR unit is integrated into a Fourier-Transform infrared spectroscopy (FTIR) setup for the infrared absorbance measurements. For the ATR-FTIR measurements, Vertex 80V spectrometers were used (Bruker Optics, Ettlingen, Germany) in combination with a liquid nitrogen-cooled mercury cadmium telluride (MCT) detector and a middle infrared (MIR) source. Device setup and spectrometer parameters for spectra acquisition were previously described in detail.^18–20^ The ATR unit (Specac Ltd., Slogh, England) was fit into the sample compartment of the FTIR instrument and aligned to an incidence angle of 45°.^19^ Measurements were conducted under constant dry airflow to prevent the sample chamber’s sharp atmospheric water vapor absorbance bands.^18–20^ As described before, multichannel measurements were performed with a motorized stage.^20^

### Functionalization of the ATR surfaces

The αSyn absorbance spectrum is revealed by difference spectroscopy between αSyn bound spectra and spectra taken before αSyn binding, eliminating the large background absorbance spectra of the surface layer and the water background.

The surface functionalization has already been described in detail.^18,19^ In this study, an improved functionalization as described in the patents WO2024003213A1 and WO2024003214A3 was used, which will be described in detail elsewhere. The improvements generally led to optimized stability and inertness of the ATR surface compared to the former publications.^18,19^ The surface functionalization was monitored by recording the respective infrared spectra in each step. This allowed precise control over each reaction step. The binding of all αSyn from the body fluid to the antibody-coated surface was conducted in a circulation mode. In contrast, subsequent washing steps in a flowthrough mode were used to rinse all unbound components present in the sample by buffer.

The capture antibody used in this study was extensively tested for binding of different αSyn-conformers covering monomeric, oligomeric, and fibrillary structures (example in **ESI Figure 3S**). ELISA and external SPR analysis provided affinity information. The antibody was developed at AC Immune SA, Switzerland. Functionalized surfaces without antibodies were used to assess the specificity for αSyn and the inertness of the functionalized surface towards excessive amounts of antigen or CSF (**Figure 1B, ESI Figure 5S**).

**Figure 3.**
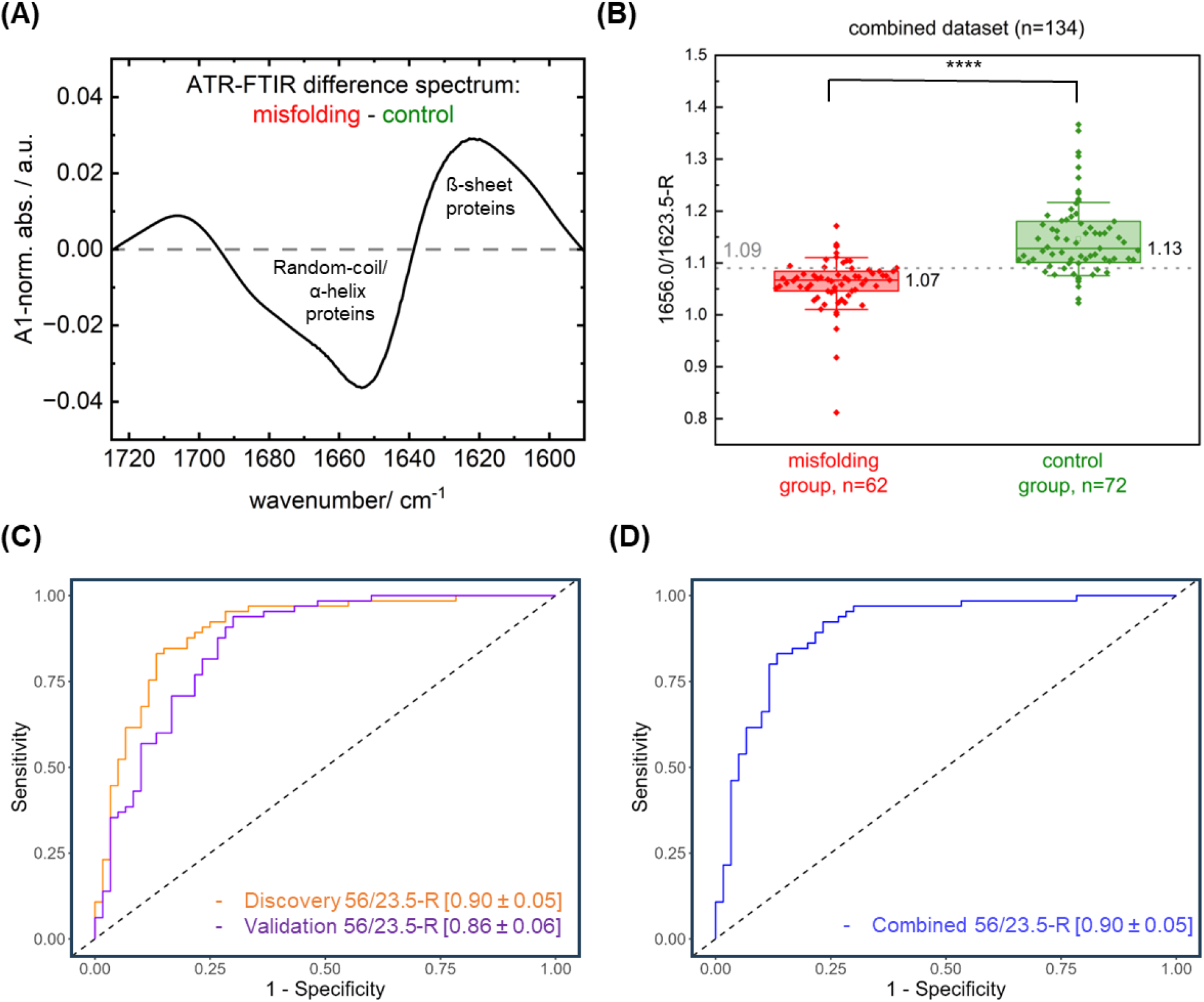
iRS analysis of αSyn secondary structure distribution in CSF. **Panel A**: The group-level changes in the Amide-I-band as normalized and zoomed difference spectra. When calculating the difference spectra (normalized misfolding (n=62) – control group (n=72)), positive absorbance values at 1623.5 cm^-1^ reflect increased β-sheet structures, while negative absorbance values at 1656.0 cm^-1^ reveal the connected decrease in α-helical/random-coil structures for PD/MSA samples compared to control samples. **Panel B**: A Boxplot of the best performing spectral feature (1656.0/1623.5-ratio) for the combined dataset, including 62 cases classified as misfolding positive (PD/MSA diagnosis) and 72 cases as controls where every point represents a single patient. A single threshold (1.093) discriminates both groups. A Mann-Witney-U test revealed statistically significant group differences with p<0.0001 (****; CI=95 %). Box and whisker plots show median value (vertical line), interquartile range (boxes), and standard deviation (whisker). **Panel C**: The ROC-AUC analyses αSyn misfolding vs. disease controls (control) in the discovery (orange) and validation study (purple) utilizing the 1656.0/1623.5 cm^-1^ spectral ratio and retrieved by a log. regression model considering age and sex. In the discovery study, an AUC of 0.90 ± 0.05 was reached, while the AUC in the validation study was at 0.86 ± 0.06. **Panel D**: The ROC curve of the combined dataset (n=134), categorized into a misfolding group (PD/MSA) and a disease control group without expected misfolding. The ROC curve was retrieved by a log. regression model considering age and sex. The AUC value is 0.90 ± 0.05 utilizing the 56/23.5-ratio. Abbreviations: αSyn, alpha-synuclein; CBD, corticobasal degeneration; CSF, cerebrospinal fluid; control, disease control; iRS, immuno-infrared-sensor; PD, Parkinson’s disease; MSA, Multiple System Atrophy; ROC-AUC, receiver operating characteristic area under curve.

Continuous spectra recording during αSyn binding allowed tracking of minute changes in aqueous background or temperature. One of four independent measurement channels was used as a reference channel by measuring a buffer instead of a sample, allowing for background monitoring and, if necessary, respective correction. After system stabilization in PBS buffer, sample background spectra were measured. CSF samples were circulated over the surface for 120 minutes without additives. After sample circulation, the system was changed to flowthrough mode for sample washing, and loosely bound materials were rinsed away with PBS buffer. Sample wash spectra were recorded and reflected the αSyn bound spectra. For efficacy testing of the setup, the supernatants after circulation mode were collected and snap-frozen for concentration determination of αSyn.

### Spectra processing and data analysis

Data analysis was performed with an in-house developed spectral software solution and scripts for MATLAB (Mathworks Inc., Natick, MA, US, MatlabR2020b). For the detailed analysis, kinetics at specific wavenumbers indicating αSyn binding, e.g., at 1550 cm^-1^ for bound protein, and whole spectra (4000-1480 cm^-1^) indicating potential artifacts were analyzed. Raw spectra were averaged and corrected for water vapor and baseline. Difference spectra were calculated between the spectra before sample measurement (sample background) and the spectra of sample wash. The difference spectra were smoothened. The MATLAB script automatically generated spectral parameters, like the absolute amide-I-peak maximum, the amide-I center of mass maximum, or spectral ratios, e.g. 1656.0/1623.5-ratio. The different read-outs reflect the overall secondary structure distribution of αSyn. The analysis of the Amide-I/Amide-II-ratio at peak maxima is used as a quality indicator, and spectra were only included in a range of 1.10-1.50. Distorted spectra with a Ratio <1.10 or >1.50 due to minor water background absorbance changes or temperature instabilities during the recording of the sample spectra were excluded from the analysis. Spectra with a S/N < 20 were also excluded from the analysis. S/N was calculated with S as the mean absorbance between 1560 and 1540 cm^-1^ and N as the root-mean-square of the absorbance values between 1800-1900 cm^-1^ of unsmoothed spectra.

### Statistical analysis

Descriptive statistics were used to summarize patient characteristics to compare the age and sex of αSyn misfolding cases and controls by chi-square and Mann-Witney U tests.

Receiver-operating characteristic (ROC) analyses, including calculation of area under the curve (AUC), were conducted for discrimination of individuals with αSyn misfolding and controls and by use of the ratio of 1656 cm^-1^ and 1623.5 cm^-1^, representing the distribution of monomeric α-helical/random-coil and β-sheet enriched protein structures. First, a logistic regression model was calculated with the disease status as the dependent variable and age, sex, and the read-out value as independent variables. The resulting model was then used to predict the probability that each person had Parkinson’s disease. The ROC Curves and associated AUC values were then calculated based on these predictions. We additionally performed the same analysis stratified by study (Discovery and Validation, see **Figure 3**). Moreover, the added value of the read-out was analyzed by comparison to age and sex model only (ESI **Figure 4S**)

**Figure 4.**
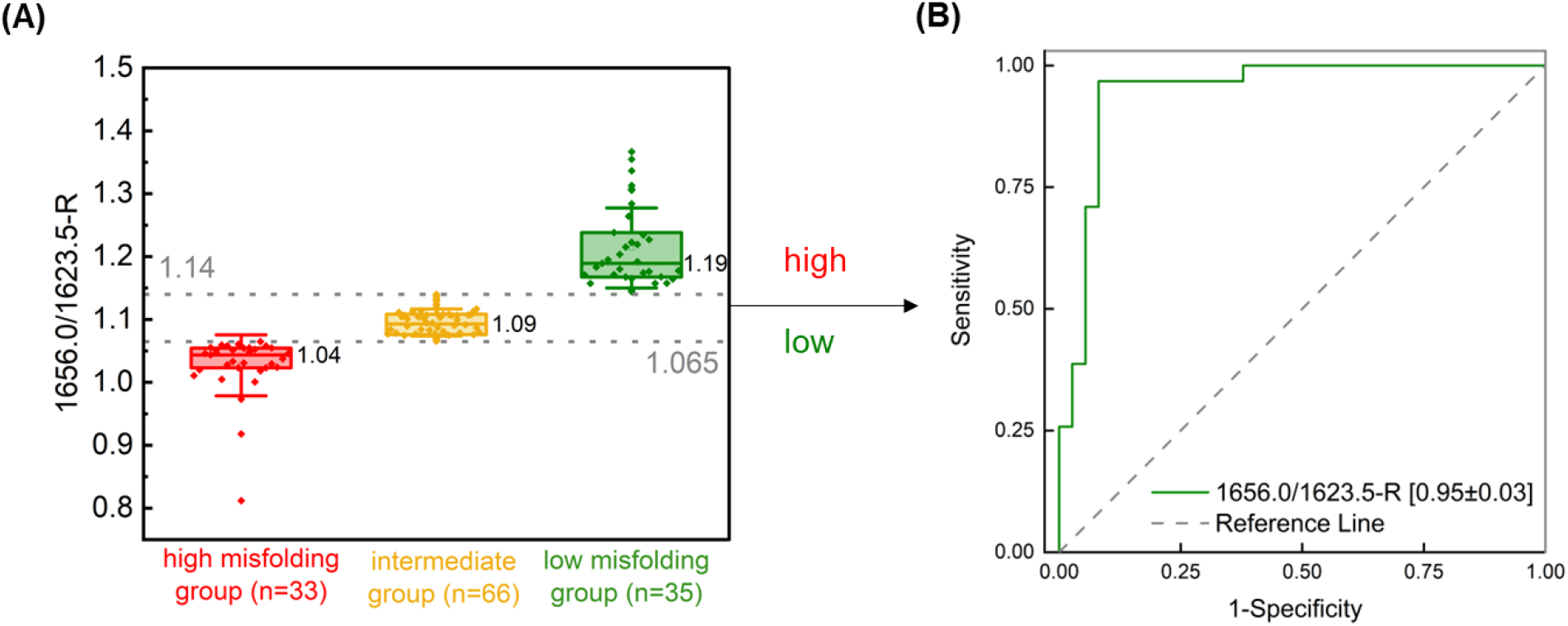
iRS double threshold for classification into high and low misfolding and an intermediate group. **Panel A:** Boxplot of the 1656.0/1623.5-ratio dependent on the misfolding status determined by the iRS-measurement and categorized into three subgroups where every diamond represents a single patient. High, intermediate, and low misfolding were determined with thresholds of 1.065 and 1.14 for the 1656.0/1623.5 spectral ratio. Box and whisker plots show median value (vertical line), interquartile range (boxes), and standard deviation (whisker). **Panel B:** The AUC performance of the high misfolding group versus the low misfolding group (n=68/134) according to the clinical diagnosis and expected misfolding status. The AUC value reached 0.95 ± 0.03 with a sensitivity of 97 % and a specificity of 92 %. Abbreviations: AUC, area under the curve; αSyn, alpha-synuclein; iRS, immuno-infrared-sensor.

All analyses were conducted two-sided at a significance level of 0.05 using OriginPro 2021b or OriginPro 2024 (OriginLab Corporation, Northampton, MA, USA) and the programming language R (version 4.2.1).

## Results

### Controls

The iRS platform technology was applied to determine the secondary structure distribution of αSyn in CSF. The iRS platform and the capture antibody’s performance were extensively characterized by the iRS platform itself and orthogonal techniques (ELISA, western blots, SPR) to qualify the analysis for CSF.

**Figure 1** shows the binding of the different αSyn conformations by the capture antibody in Panel A. In order to determine the secondary structure distribution of αSyn in body fluids, the catcher antibody must be able to bind all different conformers. The structure-sensitive Amide-I band showed its maximum at 1652 cm^-1^ in the case of the α-helical/random-coil monomers. The maximum was at 1647 cm^-1^ for dopamine-stabilized oligomers, while the most abundant oligomer species range from 700-1100 kDa.^40^ Pre-formed fibrils (PFFs) of αSyn showed a maximum at 1624 cm^-1^. The results exhibit that the used catcher antibody extracts the different conformers. The expected range of αSyn extracted by capture antibodies from CSF is from 1624 to 1652 cm^-1^. Common infrared spectroscopy wavenumber ranges for α-helices range from 1645-1662 cm^-1^, for rando-coil motifs from 1640-1645 cm^-1^, and for β-sheet motifs between 1615-1638 cm^-1^.^38,39^

The inertness of the functionalized ATR surface without the capture antibody is displayed in Panel B. Excessive amounts of the different antigen conformers, at even much higher concentrations than the physiological concentrations observed in CSF, are blocked and do not bind unspecific to the surface. Notably, the surface has enough binding capacity to bind high amounts specifically by the antibody. Furthermore, cross-reactivity of abundant proteins like HSA or amyloidogenic proteins like β-amyloid (monomers and fibrils) do not bind even in excessive amounts on the antibody functionalized surface (ESI **Figure 5S**). Importantly, the functionalized blocking layer without capture antibody does not indicate an unspecific signal from the CSF matrix either (ESI **Figure 5S**). Taken together, the results demonstrate no unspecific binding or strong cross-reactivity from CSF to the functionalized surface.

Two independent control experiments were performed to determine the extraction of αSyn by the capture antibody. In the first experiment, an already measured CSF sample was measured a second time on a freshly prepared antibody functionalized surface. **Figure 2A** shows the kinetics (left) and spectra (right) for the first and second runs. No infrared signal is observed in the second run, implying complete extraction of αSyn from CSF on the surface within the infrared sensitivity in the first run. A minimal baseline shift (2. run) occurred in the time course of measurement (6 h) was observed but did not impede the interpretation of signals. A second experiment was performed to eliminate the possibility of remaining, but not measurable, amounts of αSyn in the iRS. A commercially available quantitative ELISA for total αSyn (BioLegend, Cat. No. 448607) was used. For this, the initial CSF and the CSF supernatants after measurement were compared either on a surface functionalized with or without capture antibody (**Figure 2B**). Dilutions due to the iRS setup were adjusted. While the original CSF sample concentration was 1197 ± 37 pg/ml, the concentration in the supernatant decreased over 85 % to 161 ± 14 pg/ml, implying a high efficiency in binding αSyn from CSF. Notably, the concentration of αSyn was only slightly decreased when measuring on a functionalized surface without capture antibody (1075 ± 19 pg/ml), which agrees with no signal being observed in the iRS read-out. The samples were measured in duplicates. The results show that the functionalized surface with the catcher antibody extracts almost all αSyn from the CSF.

In summary, the functionalized surface does bind specific αSyn and does not bind other CSF compounds unspecifically. Further antibody characterization (e.g. ESI **Figure 3S**) displayed a favorable, balanced antibody binding profile with EC_50_ values in the sub-nM range.

### Discovery and validation cohorts

The iRS platform technique was applied to n=134 CSF samples from different clinical centers. The discovery study encompassed 59 individuals, with 17 out of 59 diagnosed with PD and 42 out of 59 as disease controls without signs of neurodegenerative disorders. The validation cohort comprised 75 individuals from the Paracelsus-Elena-Klinik (Kassel) and is entirely independent of the discovery cohort.

Patients with overlapping disorders were included in the validation cohort for a more challenging differential classification (details in **ESI** Table 1S). In the Kassel cohort, 40 out of 75 were diagnosed with clinical PD. Since the presence of αSyn-aggregates and conclusively αSyn-misfolding in MSA patients is known, both PD and MSA subjects were considered as misfolding positive group, while all other subjects were grouped as one disease control (control, compare ESI Table 1S) group, including CBD, FTD, and PSP subjects.^13,41,42^

The diseased group showed an average downshifted maximum of ʋ_av_PD/MSA_=1639.54 cm^-1^ compared to the control group with a maximum of ʋ_av_control_=1641.4 cm^-1^. As read-out, not the absorbance maximum itself, but a center of mass maximum (upper 80-90 % of the band) was taken, which increases robustness in small signals. A difference spectrum between all synucleinopathy (PD/MSA) and all control spectra averaged to one spectrum is performed to quantify the downshift. The difference spectrum (**Figure 3A**) between the mean control spectra and the mean misfolding spectra marks a shift from random-coil/α-helical secondary structures at 1656.0 cm^-1^ as a negative band to the β-sheet secondary structure at 1623.5 cm^-1^ ratio as a positive band. The positive/negative difference bands have comparable infrared signal integrals of 0.55 and 0.65. This shows clear-cut that the overall signal in the infrared reflects the expected transition for αSyn from random-coil/α-helical to β-sheet conformers. This misfolding is shown directly without amplification in body fluid for the first time as a native environment for αSyn in PD.

In the next step, we used the 1656.0/1623.5 ratio from **Figure 3A** as the best-performing measure instead of the downshift in wavenumbers to distinguish between controls and diseased. The ratio significantly distinguishes misfolding positive from negative cases (*p<*.*0001*, **Figure 3B**). A receiver operating characteristic area under curve (ROC-AUC) analysis with the 1656.0-1623.5-ratio is performed in the next step. In the discovery study, an area under the curve (AUC) of 0.90 (95 %-CL 0.85 – 0.96) is obtained, while in the validation study, an AUC of 0.86 (95 %-CL 0.80 - 0.93) is yielded (**Figure 3C**). Combining both datasets yielded an AUC of 0.90 (95 %-CL 0.85 - 0.96, **Figure 3D**). Notably, a logistic regression model without the iRS read-out and considering age and sex only yielded an AUC of 0.67 in this dataset (ESI **Figure 4S**), clearly demonstrating the added value of the test.

The subgroup performance was analyzed (compare **ESI Figure 6S**) in the validation cohort. Because the subgroups were still very small (≤10), the data must be confirmed in future studies by much larger numbers. Here, a first tendency is presented. The MSA patients alone could be differentiated from the control group with an AUC of 0.73 (95 %-CL 0.56 – 0.97) but worse from PD with an AUC of 0.71 (95 %-CL 0.49 – 0.93). Accordingly, a Mann-Witney-U test for the MSA and PD groups showed no significant differences (p=0.12).

We introduced two thresholds instead of one, providing three classes (**Figure 4, ESI Figure 7S**), as the misfolding negative and positive groups revealed an overlap. This reflects that the misfolding increases in a continuum between healthy controls and individuals with clinically confirmed synucleinopathies (PD/MSA), which is not accounted for by one threshold only. Individuals with a 1656.0/1623.5-ratio <1.065 were stated as a high misfolding (red) and clear synucleinopathy affliction, whereas a ratio of >1.14 reflected low misfolding (green) and no affliction. High- and low-misfolding groups depicted a significant difference (*p<*.*0001*, **Figure 4A**). The individuals in the yellow group, between the two thresholds 1.065 and 1.14, fall into the intermediate area. These individuals are not as clearly assigned as individuals of high or low misfolding groups by iRS-readout. The intermediate group reflects the misfolding continuum between a healthy and diseased state. In this line, individuals categorized into the intermediate group may hold an elevated risk for developing synucleinopathies compared to unafflicted individuals (green group) but are not yet clearly afflicted (red group). Comparing only individuals with high (red) and low (green) misfolding groups yielded an AUC of 0.95 (95 %-CL 0.89 – 1.01) with a sensitivity of 97 % and specificity of 92 % (**Figure 4B**).

## Discussion

Disease-specific fluid biomarkers for PD and other neurodegenerative diseases are urgently needed to stratify affected patients. Atypical forms of the disease or associated copathologies with partially mimicking clinical syndromes may delay the clinical diagnosis and lead to suboptimal treatments. Robust biomarkers with high sensitivity and specificity that can be assessed *intra-vitam* are urgently needed since a confident diagnosis is currently only feasible through post-mortem neuropathologic examination. As another example, an αSyn-PET tracer will represent an important option for PD diagnosis, similar to what they have for amyloid-β in AD. However, these tracers are still in early development.^5,6^

In this study, we presented the disease-related misfolding of αSyn in CSF of PD/MSA patients as a promising fluid biomarker for the stratification of Parkinson’s disease. Misfolding of αSyn has been previously linked to disease progression, neuronal cell death, and disease pathology (Lewy bodies and neurites) in numerous studies, and therefore αSyn demonstrates a promising biomarker candidate.^43^ Because misfolding is expected already in NSD stage 2, we propose that similar to misfolding of β-amyloid in AD, αSyn misfolding might also be a very early body fluid marker in prodromal or pre-motor PD for indicating at-risk status.^8,44^

Promising results, indicating that misfolding of αSyn is an excellent fluid biomarker in Parkinson’s disease, are obtained by SAA assays. They indirectly reflect the misfolding of αSyn by amplifying competent misfolded conformers with added monomeric αSyn in the SAA. This currently represents a promising PD test, mainly in CSF. The assay has reached high sensitivities and specificities (> 90 %) in CSF. However, the read-out depends on the recombinant monomeric αSyn, the competent amplified αSyn conformers, and the assay conditions.^16^ Therefore, a considerable variation in the read-out between different labs may be observed. A harmonization for future standardized applications is needed and ongoing because quantitative read-outs of different tests developed should ideally be comparable, and expansions into peripheral matrices beyond CSF are examined.^16,17^ Other quantification strategies for disease classification using reduction of αSyn concentration due to deposition in Lewy bodies could not discriminate between patients afflicted with the disease and controls.^43^

Furthermore, there is an ongoing conceptual debate about whether PD and the spectrum of Parkinsonian disorders shall be primarily regarded and differentiated as clinical diagnoses or if biomarkers and genetics should receive a much more prominent role.^7,8,45^

Utilizing the iRS-platform, the synucleinopathies PD and MSA could be distinguished from controls by misfolding of αSyn when comparing only individuals with high (red) and low (green) misfolding groups yield an AUC of 0.95 (95 %-CL 0.89 – 1.01) with a sensitivity of 97 % and specificity of 92 % (**Figure 4B, ESI Figure 7S**) or by single decisive threshold (AUC 0.90, **Figure 3**), which is similar to the accuracy from SAA. In contrast, an AUC of 0.67 is reached without the iRS read-out and only based on demographics (age, sex). This may be explained by the unbalanced gender distribution in the case of PD/MSA cases (M:F 77:23, compare **Table 1**).

An in-depth analysis of the iRS spectra directly marks the transition from α-helical/random-coil to β-sheet in CSF (**Figure 3B**). The secondary structure distribution of all different conformers of αSyn is measured with the iRS platform, whereas, in the SAA conformers, only the seeding competent conformers are amplified and provide the read-out. Oligomers with high α-helical content (compare **Figure 1**) or β-sheet content of not fibrillar extended type may be underrepresented or not detectable in the SAA assay. The SAA platform only provides a positive or negative result, whereas the iRS result offers a quantitative measure of the continuum of disease progression. Although this needs to be investigated in further studies, this benefit could potentially answer the question of whether or not patients with prodromal PD (stage 2 NSD), e.g., REM sleep behavior disorder, progress to PD or associated αSyn aggregation disease through an early risk indication in a disease continuum. So far, the inability to predict the timespan in which these patients develop PD is a significant hurdle for developing disease-modifying therapies and prevention studies.^46^

Furthermore, the stratification of PD and the consideration of potentially overlapping neurodegenerative diseases in view of a cross-disease spectrum is essential for predicting a positive therapy response and facilitating the development of individualized therapies, as already seen for cancer. Therefore, secondary structure distribution profiles of overlapping syndromes may differentiate between neurodegenerative diseases, but these will require extended subgroup studies.

Secondary structure differences exist, e.g., in PD vs MSA-fibrils as resolved by cryo-EM. However, cryo-EM conditions in a vacuum at 77 K are rather artificial and might not reflect the secondary structure under physiological conditions and temperature. We believe that the iRS technology must confirm these findings under physiological conditions.

Concerning differential diagnosis of clinically overlapping syndromes, the subcohort of MSA patients was too small for reliable statements. It was reduced even more by two cases of an initial clinical diagnosis of MSA, which were changed to NPH and PSP but showed misfolding by iRS and additional SAA-positivity (**ESI Figure 6S**).

57 % (24/42) of the individuals diagnosed with PD or MSA in the validation cohort showed a BBB dysfunction, indicated by an albumin quotient (CSF/Serum) larger than the age-dependent norm value. 19 out of 24 (79 %) showed an iRS-misfolding positive read-out. From the 18 negative cases, three were negative with the iRS, while 15 were positive for αSyn misfolding. In contrast, only 3 out of 27 controls had a BBB-dysfunction indicated; however, all were negative by iRS read-out. The correlation of disease diagnosis or misfolding-status positivity with BBB dysfunction may indirectly indicate a higher oligomeric/protofibril burden.^47^ Interestingly, 5 out of 24 of the patients with PD/MSA diagnosis and BBB-dysfunction present a negative misfolding read-out, which may be due to oligomers with less β-sheet content (compare **Figure 1**) rather than the presence of β-sheet enriched protofibrils.

The iRS platform is well-stratified with orthogonal techniques. The platform extracts >85 % of all αSyn out of CSF by the catcher antibody while unspecific binding is prevented (**ESI Figure 5S**). Only 10 % of the analyte was lost without a capture antibody (**Figure 2**). Since no iRS-signal was observed on the blocked layer, the loss may be due to the surrounding tubing or caused by not matching the dilution factor exactly, but not due to the functionalized surface itself.

All experiments are performed on home-built instruments, although precision, reproducibility, and device comparability are high (**ESI Figures 8S & 9S**). In the future, the CE-certified commercial iRS1.1 instrument (betaSENSE) will improve these parameters further.

In addition to the observed changes of target proteins, natural binding partners of αSyn in CSF may contribute to the iRS read-out, possibly enhancing it. This would be the case if binding partners of misfolded and native fold αSyn are also different. A detailed analysis of the captured proteins will be performed to characterize the possible secondary binding partners using a newly developed and iRS-integrated mass spectrometry workflow.

We propose a combined iRS analysis of β-amyloid, tau, αSyn, and other biomarkers to address obstacles of co-pathologies and patient stratification based on protein misfolding.^21^ Neuropathologic studies show that the incidence of mixed co-pathology is high among neurodegenerative disorders and modifies disease symptoms and progression.^48^ Therefore, identifying co-pathology patterns could help better understand overlapping clinical symptoms and develop targeted therapy strategies.

Since misfolding is believed to occur before the onset of clinical symptoms, early diagnosis and risk prediction using the platform are addressed in a follow-up study, including isolated RBD patients (video-supported polysomnography verified) showing an enhanced risk for conversion to synucleinopathies.^49^ For risk assessment and early stages, the double threshold classification has shown an intriguing result in the first subset of isolated RBD patients (Paracelsus-Elena-Klinik Kassel, Germany) and will be followed up.

To sum up, we developed the extension of the iRS technology from Alzheimer’s to synucleinopathies, which for the first time allowed differentiation of synucleinopathies and controls by direct measurement of αSyn misfolding as conversion from α-helical to β-sheet by difference spectra in the unmodified CSF matrix.

Further studies are planned concerning the biological classification (SynNeurGe frame network) of overlapping syndromes or copathologies in larger cohorts. Lastly, as misfolding is an early biomarker, iRS analysis of high-risk individuals (e.g., RBD) over time is strived for risk assessment and prediction value.

## Supporting information

Electronic Supporting Information

## Data Availability

The data supporting this study's findings are available from the corresponding author, K.G., upon reasonable request.

## Author contributions

M.S., L.B., J.G., C.K., L.T., B.M., and K.G. designed research. M.S., G.G., M.M., N.W., D.H., D.D., A.H., S.G., J.S., L.B. performed research. M.M., G.G., N.W., L.L, R.W., J.G., C.K., K.G. innovated the patented current surface chemistry and sensor design. M.S., R.D., S.W., E.H.K., J.G., L.T., B.M. analyzed data. M.S., L.B., and K.G. wrote the paper. M.S., L.B., S.W., E.H.K, C.K., J.G., L.T., B.M, K.G. revised the paper. All authors read and approved the final manuscript.

## Acknowledgments

AC Immune SA developed and produced the αSyn antibody used in this study. The authors greatly thank AC Immune SA for its provision and support in all antibody-related topics. The authors thank betaSENSE GmbH for providing the antibody, antibody-related protocols, and staff.

