## Supplementary material for "Alpha-synuclein misfolding as a fluid biomarker for Parkinson’s disease and synucleinopathies measured with the iRS platform": Electronic Supporting Information

### Methods

#### Thioflavin T fluorescence for antigen verification

Antigen testing was performed for every generated or purchased batch. The indication of β-sheet structures in fibrillary (or some oligomeric) antigens and the absence of those in monomeric stock material was tested by a Thioflavin T (ThT) fluorescence assay. Antigen stock material, stored at -80°C in single-use 346 µM aliquots, was thawed slowly on ice for triplicate ThT measurement.

25 µM ThT-PBS-solution (PBS: 137 mM NaCl, 7.8 mM $Na_2HPO_4$, 1.4 mM $NaH_2PO_4$, 3.2 mM KCl pH 7.4) was used for dilution of the antigen material (compare section "antigen preparation"*)* to a final concentration of 8.6 µM in any wells of the 96-well-plate (Greiner Bio-One GmbH, Frickenhausen, Germany, Greiner 96 F-Bottom fluorescence plate, cat. no. 655076) assigned for measurement. Controls were included with sample buffer instead of sample (blank), monomeric material as negative control in case oligomeric or fibrillary material testing and vice versa. The plate was incubated for 60 minutes at room temperature while shaking at *300* rpm (VWR International LLC, Radnor, US, Incubating Microplate Shaker with Lid). The read-out was performed by recording a fluorescence spectrum in the range of 410-460 nm (excitation scan, emission wavelength 490 nm) and 480-620 nm (emission scan, excitation wavelength 450 nm) using the plate reader (BMG LABTECH GmbH, Ortenberg, Germany, CLARIOstar Plus). Scan parameters included 20 flashes/well, a bandwidth of 10 nm, 300 rpm double orbital shaking for 5 s before plate reading, and a settling time of 0.5 s. The gain was adjusted at expected peak wavelengths (450/483 nm), and the focus was set to autofocus.

The presence of β-sheet rich aggregates was confirmed if the excitation scan maximum was shifted from 413 nm (free ThT) to 450 nm (intercalated ThT) and an emission increase by at least a factor of 20 was present when calculating the mean intensities at 482 nm against the blank without a sample.[1,2] A representative example of the ThT spectra and calculation of the ThT-emission factor is presented in **ESI Figure 1S**.

ATR-FTIR antigen spectra in buffer

For ThT-independent verification of the secondary structure features and estimation of α-helical content in β-sheet enriched antigens, the Bruker Alpha instrument (Bruker optics, part of Bruker Corp., Billerica, Massachusetts, US) with a single reflection diamond ATR unit was used. In contrast to the measurements on specifically functionalized silicon crystals with capture antibody, the antigens on the Bruker Alpha were directly measured in buffer against the sample buffer as background. Typically, 10 background spectra (each 30 scans) were averaged and used recalculation of the averaged sample signal (10 spectra, each 30 scans). Spectra were recorded from 4000-400 $cm^{-1}$ and a spectral resolution of 2 $cm^{-1}$. Spectra were recorded using the OPUS software and then analyzed using the in-house MATLAB script to extract spectral features. The absolute Amide-I band maximum and spectral ratios were used to estimate β-sheet enriched structures compared to α-helical/random-coil monomers (compare **ESI Figure 1S**).

Antibody production

The native antibody was produced in large batches in a CHO cell line at AC Immune, purified, tested for affinity towards aS-M and aS-PFFs in internal Elisa setups and SPR, and provided for measurements by dry-ice shipping. Antibody was stored in aliquots at -80 °C in PBS buffer until usage. All CSF study samples were measured with the same antibody batch.

Antibody-labeling and attachment to surface

A DBCO-containing molecule was attached to the antibody according to published protocols for covalent attachment to the functionalized surface.[3,4] The labeled antibody was compared to the native antibody in an indirect ELISA for $EC_{50}$ determination and on the sensor surface for antigen capture for quality control (compare **ESI Figure 3S**).

Indirect ELISA for $EC_{50}$ determination of (labeled) capture antibody molecules

The following protocol of an indirect ELISA for $EC_{50}$ value determination was used to assess the functionality of the distinct antibody batches before and after labeling with DBCO next to ATR-FTIR measurements. Next to this, the binding of antigens to the antibody was characterized externally by SPR and Elisa (AC Immune).

For the ELISA, the 96-well Nunc-Plate (Thermo Fisher Scientific Inc., Waltham, US, cat. no. 439454) was coated with 1.5 ug/ml αSyn-M or αSyn-PFF (Stressmarq Bioscience Inc., Victoria, Canada, cat. no. SPR-321 and SPR-322) in 50 µl coating buffer (0.1 M $Na_2CO_3$, 0.1 M $NaHCO_3$, pH 9.7) for 1 h at 300 rpm shaking (Thermo Fisher Scientific Inc., Waltham, US, Wellwash™ Versa). The plate was washed thrice (0.1 M Tris, 0.15 M NaCl, 0.05 % Tween20, pH 7.4), and wells were blocked with 1% skim milk in the wash buffer for 30 minutes. After repeated washing, the antibody to test was applied by a serial dilution in PBS starting from 750 nM for 1h at 300 rpm shaking. After repeated washing, wells were incubated with the secondary antibody (anti-mouse IgG-HRP, Merck KGaA, Darmstadt, Germany, art. no. A3673) for 1 h. Lastly, and after washing, the substrate (Sigma-Aldrich Co. LLC, St. Louis, Missouri, US, art. no. P9187, Sigmafast™ OPD tablet) was automatically injected by needle (BMG LABTECH GmbH, Ortenberg, Germany, CLARIOstar Plus).

Absorbances were measured at 450 nm and 620 nm (background subtraction). For $EC_{50}$-determination, raw absorbances were blank and corrected using MARS software (BMG LABTECH GmbH, Ortenberg, Germany). After that, background absorbance values at 620 nm were subtracted from blank corrected values at 450 nm. The difference signal was used to calculate the $EC_{50}$ values with a software-embedded 4-parameter logistic fit model. An example of signal curves and calculated $EC_{50}$-values is presented in **ESI Figure 3S**.

#### Colorimetric aS quantification in CSF by LEGEND MAX™ ELISA

For quantification of aS in original samples and measurement supernatants, the colorimetric kit LEGEND MAX™ Human α-synuclein from BioLegend® (San Diego, California, US, cat. no. 448607) was utilized with a reported sensitivity of 1.80 ± 0.21 pg/ml and a standard range of 10.2-650 pg/ml. All steps were conducted as stated in the manufacturer's protocol. CSF original samples were diluted 1:10 to match the standard range. Meanwhile, supernatants with a predilution of 1:3 due to ATR-FTIR measurement were adjusted to fit the original sample dilution before plate application. Samples and calibration curves were run in duplicates. Washing of the plate was programmed with the plate washer Wellwash™ Versa using 4x washing with each 300 µl/well. Absorbances at 450 and 570 nm (background) were read using the CLARIOstar Plus (BMG LABTECH GmbH, Ortenberg, Germany).

Plate reader settings included 22 flashes/well/cycle and two excitations at 450 and 570 nm measured after adding the stop solution. 300 rpm double orbital shaking for 5 seconds was done before read-out. The data was analyzed in the MARS software (BMG LABTECH GmbH, Ortenberg, Germany) by blank correction, background subtraction (450-570 nm), and a 4-parameter logistic fit model, as described before. The standard curves were used to calculate the absolute αSyn concentrations in original samples and supernatants after measurement on a capture-antibody coated surface (=efficiency of setup) and on a surface without capture antibody (=system loss), as demonstrated in **Figure 2**.

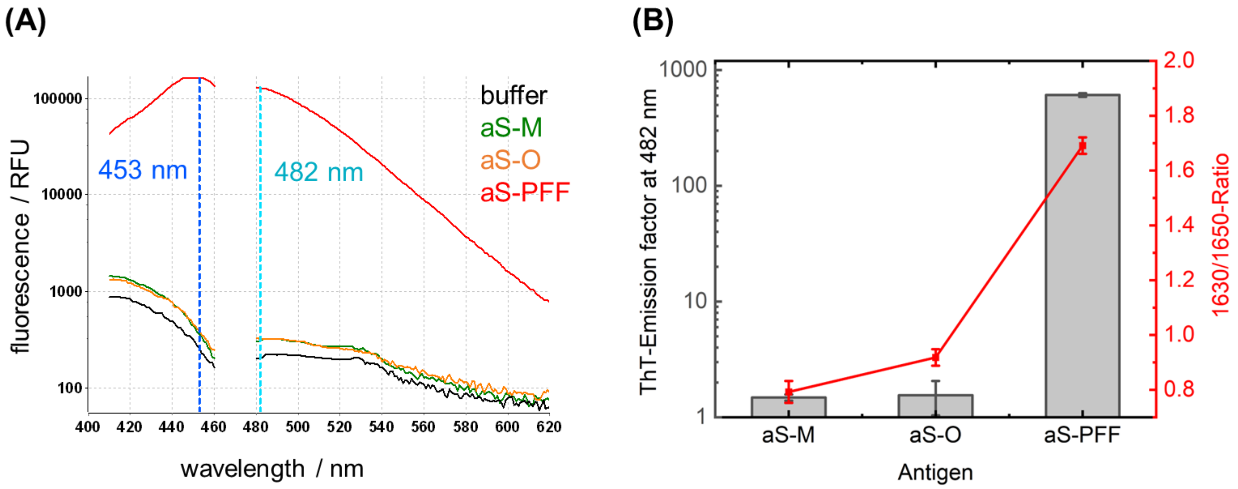


**Figure 1S**: **Panel A:** The mean ThT-excitation and emission spectra from sample triplicates, recorded from 410-460 nm and 480-620 nm and displayed in relative fluorescence units (RFUs). For this, triplicates of αSyn-M (Stressmarq Bioscience Inc., SPR-321), αSyn-O (Stressmarq Bioscience Inc., SPR-466), and αSyn-PFFs (Stressmarq Bioscience Inc., SPR-322) were prepared with a final concentration of 7.14 µM in 25 µM ThT-PBS buffer, incubated for 60 minutes at ambient temperature in the 96 F-bottom fluorescence plate and measured with the plate reader in spectral scan mode. The excitation scan was performed from 410-460 nm, while the emission scan was performed from 480-620 nm, with a bandwidth of 10 nm and 20 flashes/well. The gain was adjusted at expected maxima (453/482 nm), and autofocus obtained focal height. Before read-out, the plate was shaken at 300 *rpm* for 5 seconds. **Panel B:** The ThT-emission factor was calculated with mean intensities at 482 nm by division of mean intensities of the buffer at 482 nm (25 uM ThT-PBS). Moreover, the 1630/1650-R, derived from ATR-FTIR measurement in duplicates, was plotted against the ThT factor since the increased 1630/1650-R shows increased ß-sheet content of the given protein and decreased α-helical/random-coil content. To obtain the 1630/1650-R without the influence of binding tendencies of antibodies, antigens were measured directly on the Bruker alpha ATR-FTIR cell against the respective buffer (PBS, pH 7.4). The 1650/1630-R was retrieved automatically by the same MATLAB script used for the CSF samples after averaging 10 sample spectra for increased S/N.

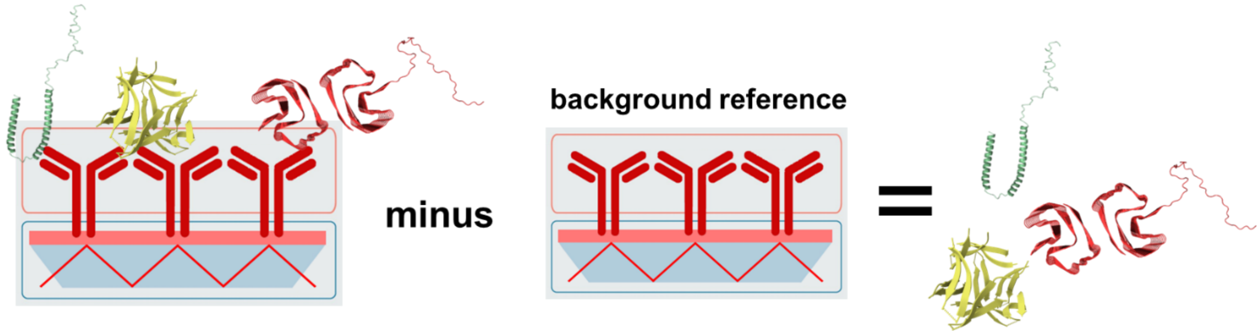


**Figure 2S:** Concept of the iRS. Under total reflection conditions, the infrared beam is guided through an ATR crystal (in blue). The evanescent wave measures the absorbance in a 500 nm thin layer on the surface. A blocking layer (red) prevents unspecific off-target binding while a covalently bound antibody captures the target molecules, in this case, αSyn.[5–8] A background reference spectrum is taken before sample application and subtracted as background reference. The difference spectrum reveals the absorbance spectrum of all bound conformers only. Depending on the distributions of the conformers, the read-out varies between 1650 $cm^{-1}$ wavenumbers for a pure α-helical/random coil distribution and 1630 $cm^{-1}$ for a pure β-sheet conformer distribution. Mixtures absorb in between depending on the conformer distribution. The downshift of the read-out represents the disease progression in a continuum.

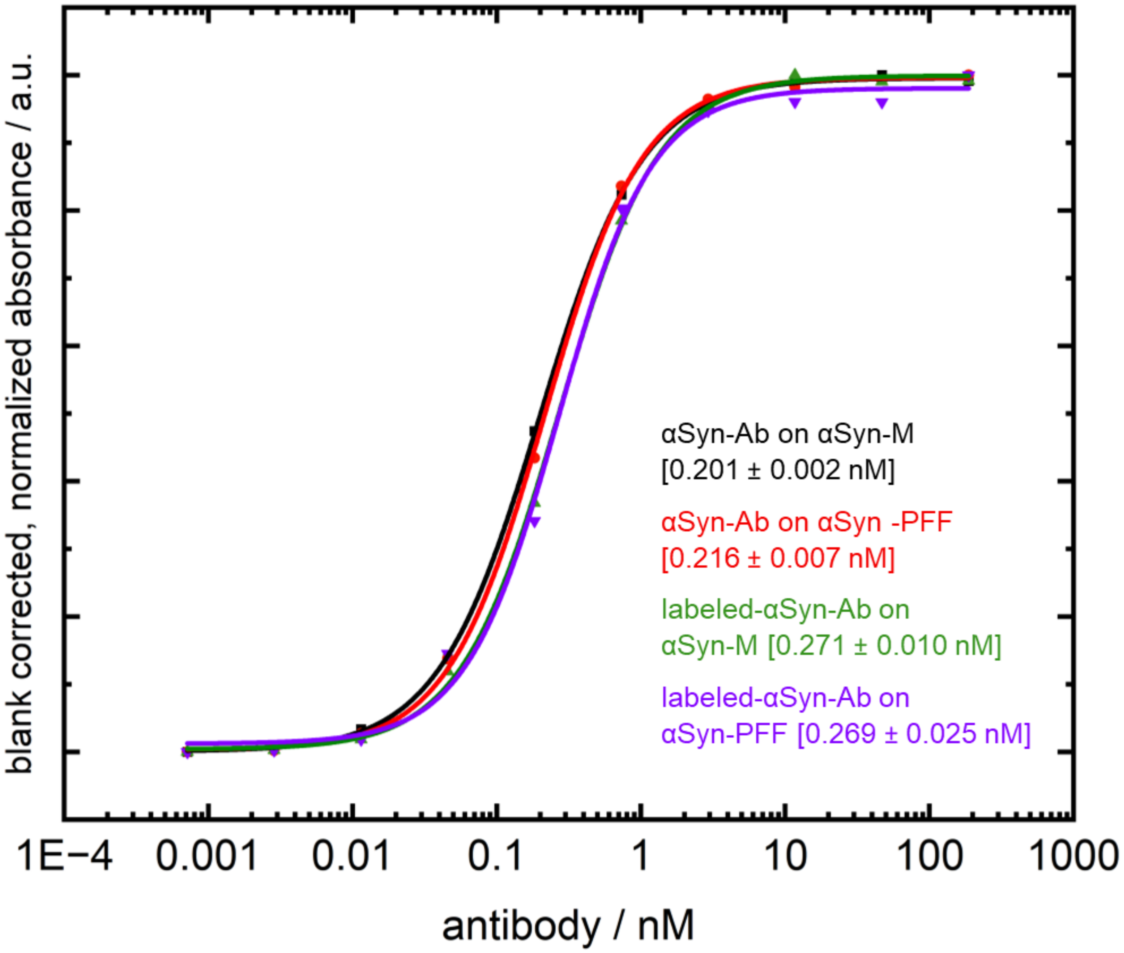


**Figure 3S**: Indirect ELISA for $EC_{50}$-value determination of native and labeled capture antibody according to given protocol (ESI Materials). $EC_{50}$-values were determined from blank corrected absorbance difference at 450 - 620nm using a 4-parameter logistic fit model on normalized data. Values demonstrate the fit results of the native and labeled antibody towards αSyn-M (Stressmarq Bioscience INC. SPR-321) and αSyn-PFF (Stressmarq Bioscience INC. SPR-322), as used in the ThT and ATR-FTIR experiments. $EC_{50}$ values from the 4-parameter logistic fit model are 0.20 ± 0.002 nM and 0.22 ± 0.007 nM for native antibody, while labeled antibody shows $EC_{50}$ values of 0.27 ± 0.010 nM and 0.27 ± 0.025 nM.

| | Demographics | | | Demographics + iRS | | |
|---|---|---|---|---|---|---|
| **Characteristic** | **OR**[1] | **95% CI**[1] | **p-value** | **OR**[1] | **95% CI**[1] | **p-value** |
| Age | 1.020 | 0.988, 1.055 | 0.2 | 1.027 | 0.986, 1.076 | 0.2 |
| Gender | | | | | | |
| m | — | — | | — | — | |
| w | 0.264 | 0.119, 0.564 | <0.001 | 0.298 | 0.105, 0.817 | 0.020 |
| readout10 | | | | 0.030 | 0.006, 0.101 | <0.001 |

[1]OR = Odds Ratio, CI = Confidence Interval

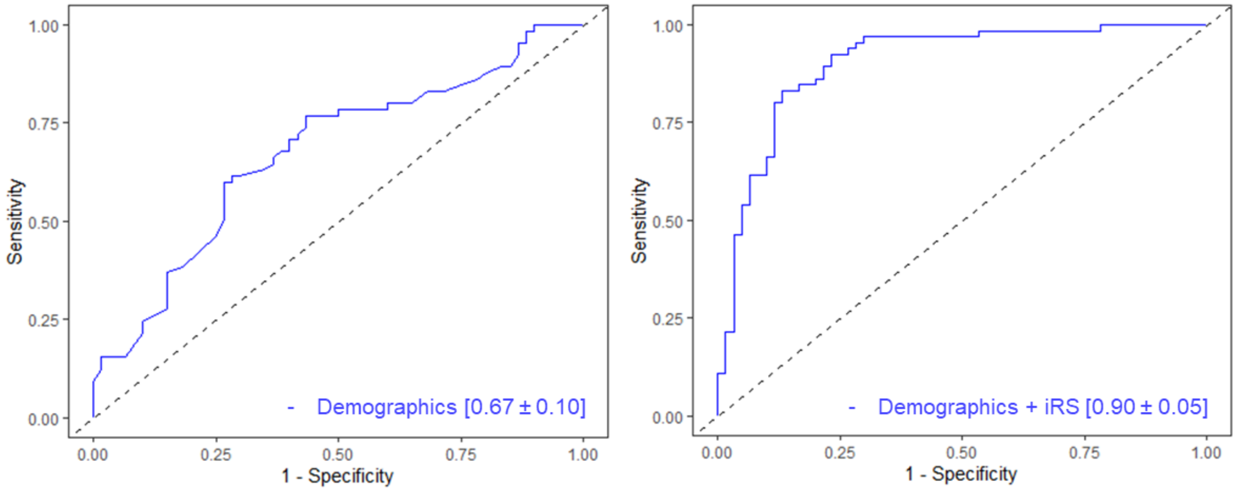


**Figure 4S**: Logistic regression models for the combined dataset. The first model, "Demographics" accounts for age and sex only, while the second model "Demographics + iRS," also accounts for the iRS assay read-out. The clinical diagnosis is always used as the dependent variable. Calculated models with characteristics in the upper table were utilized to calculate ROC-AUC curves for both models. The demographics-only model showed an AUC of 0.67 ± 0.10, while with additional iRS read-out, the AUC increased to 0.90 ± 0.05.

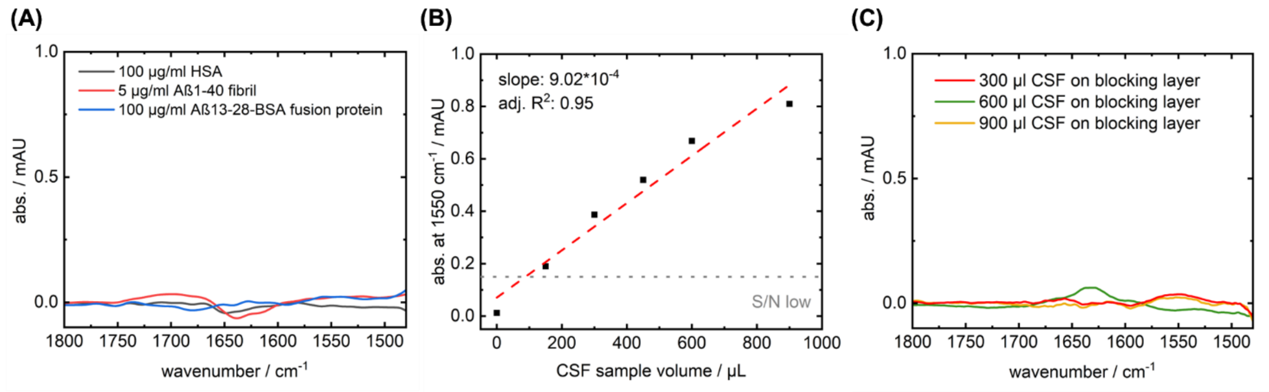


**Figure 5S**: **Panel A**: The cross-reactivity test by mean wash spectra in PBS on an αSyn -capture antibody surface with HSA, $Aß_{1-40}$-fibrils, and $Aß_{13-28}$-BSA fusion protein (produced by two-step sulfo-nhs reaction with primary amines; Thermo Fisher Scientific™, art. no. 39269) in 5-100 µg range in 1 ml total volume. No bound signal is observed in all cases; thus, no cross-reactivity to abundant HSA and other amyloidogenic proteins such as β-amyloid in the form of a $ß\text{-amyloid}_{1-40}$ fibril (preparation according to Shin et al.) or conjugated $β\text{-amyloid}_{13-28}$ (monomeric form) in high concentrations is observed.[9] **Panel B:** The amount of immobilized protein on an αSyn capture antibody surface by absorbance at 1550 $cm^{-1}$ in relation to the used CSF volume. Since S/N is critical when determining the secondary structure sensitive amide-I spectral features and samples vary naturally, volumes of 300 µl were analyzed. The assay signal between 300-900 µl shows good alignment to linear fit, implying enough capture antibody molecules for the target analyte as expected for antibody excess surfaces. **Panel C:** The inertness of the blocking layer without the capture antibody on the surface for 300-900 µl CSF sample volume as mean wash spectra in PBS. No specific signals were observed at 1550 $cm^{-1}$ (=bound protein) or in the amide-I region (1600-1700 $cm^{-1}$) without the capture antibody on the assay's blocking layer.

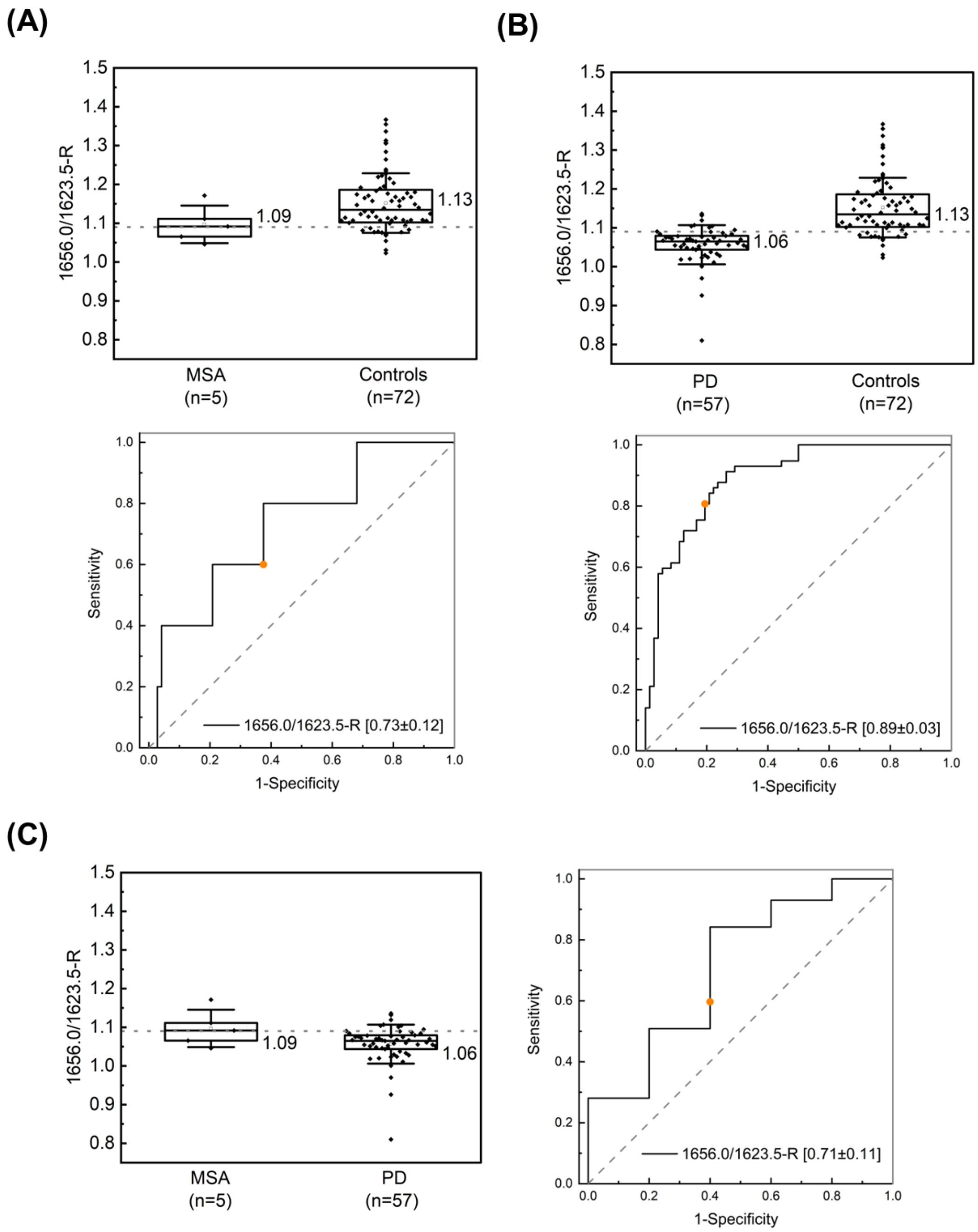


**Figure 6S**: ROC-analysis for subgroups. **Panel A**: MSA versus control subjects display an AUC of 0.73 ± 0.12. Thus, the five presented MSA cases are closer to controls than PD subjects (AUC 0.89 ± 0.03, **Panel B**) regarding the iRS-readout. The separation between the two synucleinopathies, MSA and PD, is the worst of all subgroups with an AUC of 0.71 ± 0.11 (**Panel C**), but still comparable to MSA vs control separation. The whiskers of Boxplots depict 1x SD, while the given values report the median value of the groups. Note: small and unbalanced datasets in the case of MSA hinder robust interpretation of group differences and only present tendencies in affected comparisons.

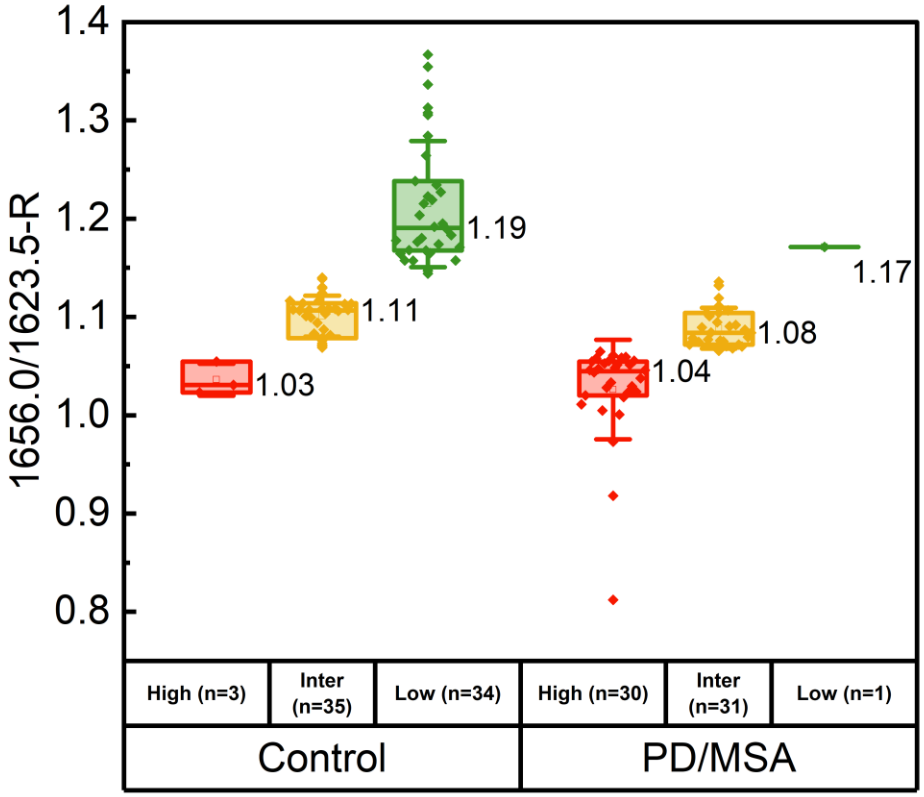


**Figure 7S**: Classification into the high, intermediate, and low misfolding group and separately shown for the control group (n=72) and PD/MSA group (n=62) in comparison to Figure 4 (main). Within the control group and according to the selected thresholds (1.065/1.14), 3 individuals fall into the high misfolding (false positive), 35 into the intermediate area (median 1.11), and 34 into the low misfolding group. In contrast, in the PD/MSA group, 30 individuals fall into the high misfolding, 31 into the intermediate group (median 1.08), and 1 (false negative) into the low misfolding group. Box and whisker plots include the median value (vertical line), interquartile range (boxes), and standard deviation (whisker).

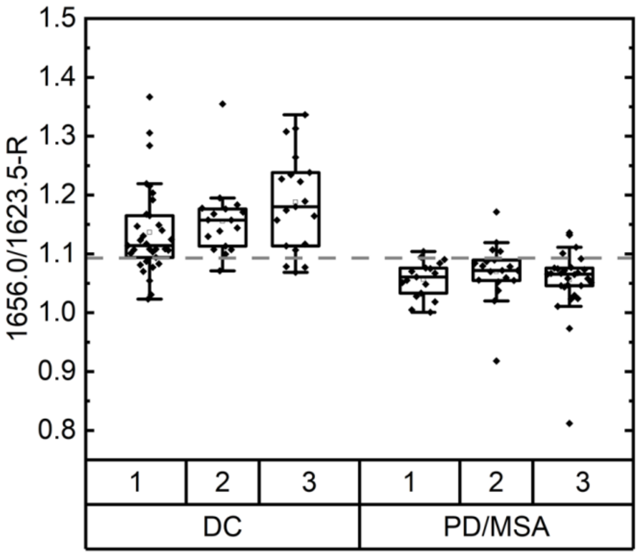


| M.-W. U Test | PM1 (n=17) | PM2 (n=19) | PM3 (n=29) |
|---|---|---|---|
| **DC1 (n=33)** | *p*<.0001 (****) | *p*<.00018 | (****) |
| **DC2 (n=17)** | (****) | (****) | (****) |
| **DC3 (n=19)** | (****) | (****) | (****) |

**Figure 8S**: Sample distribution (n=134) on three independent iRS instruments (1,2,3). Different devices with randomized sample sets are comparable, and separation is highly significant. All subgroup comparisons by Mann-Whitney U testing are listed in the adjacent table (*p*<.00018 to *p*<.0001).

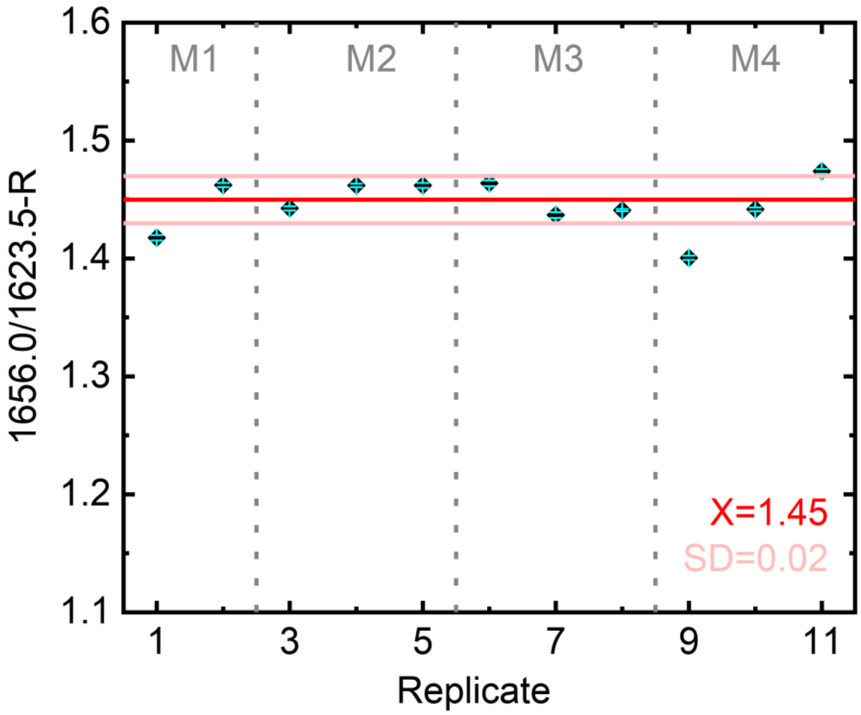


**Figure 9S**: Reproducibility measures with a pooled control CSF sample measured on the αSyn capture antibody surface. The eleven replicates were measured in four measurements (M1-M4). Routine spectra processing (WV and baseline correction, averaging spectra and reference channel subtraction, Fourier self-deconvolution (FSD), and smoothing) was performed. Each replicate is depicted with an error bar (cyan) obtained from applying a smoothing variation by the in-house MATLAB script for data analysis. The mean 1656.0/1623.5-ratio of all measurements was $\bar{X}$=1.45 with a standard deviation of $SD$=0.02.

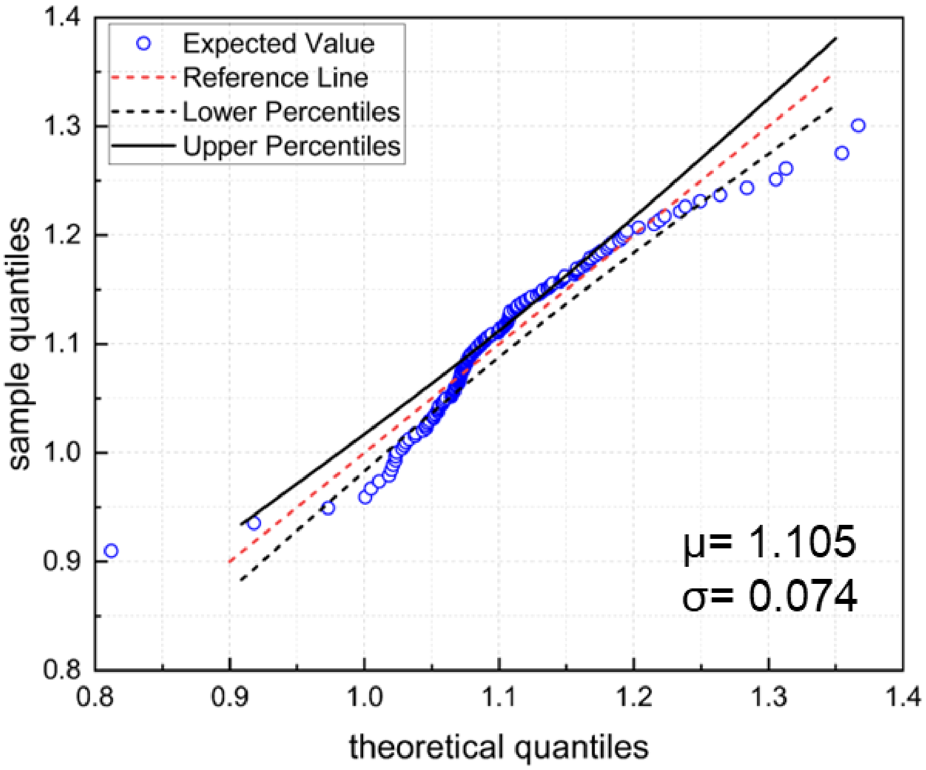


**Figure 10S**: Dataset distribution and conformity to normal distribution by normal Q-Q-Plot of 1656.0/1623.5-Ratios. The mean of the normal distribution is $\mu$=1.105 with a standard deviation of $\sigma$=0.074 for the normal distribution. Lower and upper percentiles (black line and dashed black line) mark 95 %-confidence bands, while the red dashed line shows the reference line. Since a considerable amount of data points of the combined data set (n=134) are not located within the confidence borders, the dataset is not normally distributed. Thus, non-parametric models were applied to test the significance of group separations.

**Table 1S**: Sample details of study subjects. Cohort (Discovery D, Validation V), superordinate group (Control/Synucleinopathy), diagnosis according to ICD-10, iRS single threshold classification (positive (pos), negative (neg)), iRS classification into three groups (low-, intermediate, high misfolding) and additional information.

| Sample no. | cohort | Group | Diagnosis acc. to ICD-10 | iRS-test result (1.093) | iRS-test three groups (1.065, 1.14) | Additional information |
|---|---|---|---|---|---|---|
| 1 | D | Control | Stroke, AD | neg | intermediate | N/A |
| 2 | D | Control | Borreliosis | neg | low | N/A |
| 3 | D | Control | Muscle cramps | neg | low | N/A |
| 4 | D | Control | Facial nerve palsy | neg | low | N/A |
| 5 | D | Control | PNP | neg | intermediate | N/A |
| 6 | D | Control | PNP | neg | | N/A |
| 7 | D | Control | Multifactorial gait disorder, PD, seizure | neg | intermediate | N/A |
| 8 | D | Control | MS | neg | low | N/A |
| 9 | D | Control | Seizure, subdural hematoma | pos | Intermediate | N/A |
| 10 | D | Control | NPH | pos | intermediate | N/A |
| 11 | D | Control | Intracranial hypertension | pos | Intermediate | N/A |
| 12 | D | Control | NPH | neg | low | N/A |
| 13 | D | Control | NPH | neg | low | N/A |
| 14 | D | Control | NPH | pos | high | N/A |
| 15 | D | Control | NPH | neg | intermediate | N/A |
| 16 | D | Control | Exclusion of meningitis, exitus due to cardiogenic shock | neg | intermediate | N/A |
| 17 | D | Control | Epileptic seizure | neg | intermediate | N/A |
| 18 | D | Control | Myopathy | neg | high | N/A |
| 19 | D | Control | NPH, PD | pos | low | N/A |
| 20 | D | Control | N/A | neg | intermediate | no evidence of degenerative dementia |
| 21 | D | Control | N/A | neg | low | no evidence of degenerative dementia |
| 22 | D | Control | N/A | neg | low | no evidence of degenerative dementia |
| 23 | D | Control | N/A | neg | low | no evidence of degenerative dementia |
| 24 | D | Control | N/A | neg | low | no evidence of degenerative dementia |
| 25 | D | Control | N/A | neg | low | no evidence of degenerative dementia, Psychomotor deceleration, and word-finding disorder |
| 26 | D | Control | N/A | neg | low | early phase neurodegeneration |
| 27 | D | Control | N/A | neg | low | early phase neurodegeneration, suspected vasculitis |
| 28 | D | Control | N/A | pos | intermediate | no evidence of degenerative dementia |

| | | | | | | |
|---|---|---|---|---|---|---|
| 29 | D | Control | N/A | neg | intermediate | N/A |
| 30 | D | Control | N/A | pos | high | N/A |
| 31 | D | Control | N/A | pos | intermediate | N/A |
| 32 | D | Control | N/A | neg | intermediate | N/A |
| 33 | D | Control | N/A | neg | intermediate | N/A |
| 34 | D | Control | N/A | neg | intermediate | N/A |
| 35 | D | Control | N/A | neg | low | N/A |
| 36 | D | Control | N/A | neg | low | N/A |
| 37 | D | Control | N/A | neg | intermediate | N/A |
| 38 | D | Control | N/A | neg | intermediate | N/A |
| 39 | D | Control | N/A | neg | intermediate | N/A |
| 40 | D | Control | N/A | neg | low | N/A |
| 41 | D | Control | N/A | neg | low | N/A |
| 42 | D | Control | N/A | pos | intermediate | N/A |
| 43 | V | Control | PSP | neg | intermediate | G23.1, R47.1, I10.00, I25.22, G47.39, M54.10, M43.17, M51.2, M48.09 |
| 44 | V | Control | CBD | neg | low | G23.8, G21.4, I67.3, F03, F06.3, N39.42 |
| 45 | V | Control | PSP | neg | low | G23.1, R47.1, I10.00, T88.7, R13.9 |
| 46 | V | Control | CBD | pos | intermediate | G23.8, M54.16, N40, F06.7 |
| 47 | V | Control | FTD | neg | intermediate | G23.9, G31.0, G47.9, I67.88, I10.00, E78.0, E53.8, G47.31, I44.0 |
| 48 | V | Control | PSP | neg | low | G23.1, I10.00, E53.8 |
| 49 | V | Control | PSP | neg | low | G23.1, I10.00, Z95.0, I25.29, G47.39, R13.9, R47.1 |
| 50 | V | Control | PSP | neg | low | G23.1, I25.29, E03.9, I10.00, J45.9, E78.0 |
| 51 | V | Control | PSP | neg | low | G23.1, R47.1, R48.2, K21.0, K27.7, S68.1, F32.0, E53.8, E78.0, F32.1, F06.3, F41.0 |
| 52 | V | Control | CBD | neg | intermediate | G31.0, F03, Z96.64, N40, Z92.9 |
| 53 | V | Control | CBD | neg | intermediate | G23.9, E11.90, G91.29, G25.81, E03.9 |
| 54 | V | Control | Dementia with AD, late onset | neg | intermediate | G30.1+, I10.90, R52.2, D52.9 |
| 55 | V | Control | PSP | neg | low | G23.1, G31.0+, G24.9, I67.9, Z99.3 |
| 56 | V | Control | AD, FTD | pos | intermediate | G30.9+, G21.1, F45.41, Z92.4, I10.00, I44.3 |
| 57 | V | Control | CBD with aphasia and apraxia | neg | low | G23.8, R47.1, I11.00, G47.30, E03.8, H93.1 |
| 58 | V | Control | CBD, primary progressive aphasia, tauopathy | neg | intermediate | G23.8, I10.00, M79.09, H40.1 |

| | | | | | | |
|---|---|---|---|---|---|---|
| 59 | V | Control | Dementia with AD | neg | intermediate | G30.1 |
| 60 | V | Control | AD | neg | low | F00.1 |
| 61 | V | Control | Dementia with AD, late onset | pos | intermediate | G30.1, F00.1*, E03.9, I10.90, E11.90, E53.8 |
| 62 | V | Control | Coxarthrosis, syncope, bradycardia | neg | low | M16.1; R55; R00.1; I10.0; E78.2; E03.9; D35.2; G56.0; K57.9 |
| 63 | V | Control | Hypothyroidism, carpal tunnel syndrome, glaucoma, hypertension | neg | low | E03.9; G56.0; H40.9; I10.0; E78.2 |
| 64 | V | Control | concussion, vertigo, depression, coxarthrosis | neg | low | S06.0; R42; F32.9; M16.1; F06.7; E78.0; M19.9 |
| 65 | V | Control | Constipation, cerebral transient ischaemia, nocturia | neg | intermediate | K59.0; G45.9; R35.2; E78.0; E03.9; N40 |
| 66 | V | Control | Gonarthrosis, coxarthrosis, prostatic hyperplasia | neg | low | M17.1; M16.1 ; N40; M19.9; E78.0; J45.0; M31.9; K21.0 |
| 67 | V | Control | Gonarthrosis, hypertension, coronary heart disease | neg | low | M17.1; I10.0; KHK; M19.9; L40.9; J45.0; N39.3; J32.9; M54.5; K21.0; E11.9 |
| 68 | V | Control | Hypertension, pure hypercholesterol aemia, gout | neg | low | I10.0, E78.0, M10.9 |
| 69 | V | Control | Chronic ischaemic heart disease, hypertension, diverticular disease of the intestine | neg | low | I25.9; I10.0; K57.9 |
| 70 | V | Control | PSP | pos | intermediate | G23.2, F06.3, N31.9, R47.1, R13.9, G24.0, T88.7, G47.39, G47.0, Z95.4, I§5.1, I69.4, I48.9, Z92.1, D47.3, N48.1, Z98.8, R22.9, N39.0, G62.9 |
| 71 | V | Control | NPH | pos | intermediate | G23.2, R47.1, G47.3, G25.81, G47.8, I48.0, Z92.1, M10.99, I10.00, Z96.64, Z96.65 |
| 72 | V | Control | PSP | pos | intermediate | G23.1, F02.8, S01.9, T88.7, F63.8, G24.5, G47.9 |
| 73 | D | Synucleinopathy | PD | pos | intermediate | N/A |
| 74 | D | Synucleinopathy | PD | neg | intermediate | N/A |
| 75 | D | Synucleinopathy | PD | pos | high | N/A |
| 76 | D | Synucleinopathy | PD | pos | intermediate | N/A |
| 77 | D | Synucleinopathy | PD | pos | intermediate | N/A |
| 78 | D | Synucleinopathy | PD | pos | intermediate | N/A |
| 79 | D | Synucleinopathy | PD | pos | high | N/A |
| 80 | D | Synucleinopathy | PD | pos | high | N/A |
| 81 | D | Synucleinopathy | PD | pos | high | N/A |
| 82 | D | Synucleinopathy | PD | neg | intermediate | N/A |
| 83 | D | Synucleinopathy | PD | pos | high | N/A |
| 84 | D | Synucleinopathy | PD/LBD or PD/AD copathology | pos | intermediate | N/A |

| | | | | | | |
|---|---|---|---|---|---|---|
| 85 | D | Synucleinopathy | PD | pos | high | N/A |
| 86 | D | Synucleinopathy | PD | pos | high | N/A |
| 87 | D | Synucleinopathy | PD | pos | high | N/A |
| 88 | D | Synucleinopathy | PD | pos | intermediate | N/A |
| 89 | D | Synucleinopathy | PD | pos | high | N/A |
| 90 | V | Synucleinopathy | PD | pos | intermediate | G20.10, F02.3, I48.9, Z92.1, I25.13, M72.0, E53.8, I10.00 |
| 91 | V | Synucleinopathy | MSA-P | neg | low | G23.2, R47.1, F06.3, E06.3, N39.0, B96.2, E78.0, G47.0, G47.39, G47.9, I95.1 |
| 92 | V | Synucleinopathy | PD | neg | intermediate | G20.11, R47.1, G25.81, I10.00, L73.9, R44.0 |
| 93 | V | Synucleinopathy | PD | neg | intermediate | G20.11, F02.3, F06.0, M81.99, E04.1, S22.32, E87.6 |
| 94 | V | Synucleinopathy | PD | pos | high | G20.10, R47.1, F10.6, F10.2, F06.0, E53.8, I42.0, Z95.0 |
| 95 | V | Synucleinopathy | PD | pos | high | G20.10, F44.4, F33.1, K59.0, G62.88, E53.8 |
| 96 | V | Synucleinopathy | PD | pos | high | G20.10, F02.3, M47.86, K59.0, F06.0 |
| 97 | V | Synucleinopathy | PD | pos | high | G20.11, G25.4, K59.0, I10.00, I95.1, E14.9, E78.0, I12.00, G62.9, G57.3, N40, E66.99, D47.2 |
| 98 | V | Synucleinopathy | PD | pos | high | G20.11, F02.3, F06.0, G25.4, F06.3, F41.0, N39.42, K59.0, F63.9 |
| 99 | V | Synucleinopathy | PD | pos | high | G20.11, E53.8 |
| 100 | V | Synucleinopathy | PD | pos | intermediate | G20.21, F02.3, I67.88, F06.3, G62.9, Z96.64, S46.1, I10.00, F06.0, E53.8 |
| 101 | V | Synucleinopathy | PD | pos | high | G20.11, F02.3, F06.0, E11.90, I87.20, N39.42 |
| 102 | V | Synucleinopathy | PD | pos | high | G20.11, F02.3, F06.0, G25.4, E05.1 |
| 103 | V | Synucleinopathy | PD | pos | intermediate | G20.00, F02.3, I10.00, E03.9, H40.9, I44.0 |
| 104 | V | Synucleinopathy | MSA-P | pos | intermediate | G23.2, F06.3, G47.0, E03.9, H91.2, K31.88, R94.3 |
| 105 | V | Synucleinopathy | PD | pos | high | G20.11, I10.00, I25.29, E11.90 |
| 106 | V | Synucleinopathy | PD | neg | intermediate | G20.10, F01.3, I67.3, E14.40, E11.90, R26.8, I10.00, E53.8 |
| 107 | V | Synucleinopathy | PD | pos | high | G20.10, F02.3, E04.9, I49.8 |
| 108 | V | Synucleinopathy | PD | pos | high | G20.10, F02.3, I10.00, I35.0, E04.1, E78.0, I44.0 |
| 109 | V | Synucleinopathy | PD, NPH | pos | intermediate | G20.10, G91.20, G14, F32.0, I25.29, R73.0, I70.29, Z22.3, U80.00, Z29.0, R31 |
| 110 | V | Synucleinopathy | PD | pos | intermediate | G20.00, Z85.6, G62.9 |
| 111 | V | Synucleinopathy | PD | pos | intermediate | G20.10, G25.81, I10.00, I48.0, Z92.1, M48.06, M43.16, A68.1 |
| 112 | V | Synucleinopathy | PD | neg | intermediate | G20.10, F02.3, F06.0, G25.81, G47.0, I10.00, I67.3 |

| | | | | | | |
|---|---|---|---|---|---|---|
| 113 | V | Synucleinopathy | PD, LBD | pos | high | G20.20, F02.3, G40.2, I48.2, Z92.1, I10.00, Z72.0 |
| 114 | V | Synucleinopathy | PD | pos | intermediate | G20.11, I25.12, E78.2, G62.9, N18.9, F06.3 |
| 115 | V | Synucleinopathy | PD | pos | high | G20.10, G31.82, F02.3, F02.3, F06.0, M77.3, 67.9, G47.9 |
| 116 | V | Synucleinopathy | PD | pos | high | G20.11, G25.4, G47.0, G25.81, G47.9, E53.8, G47.31, R13.9, F06.7 |
| 117 | V | Synucleinopathy | PD | neg | intermediate | G20.10, F02.3, I67.3, I10.00, E78.0, G62.9 |
| 118 | V | Synucleinopathy | PD | neg | intermediate | G20.10, G47.0, I25.13, I25.15, I10.00, E11.90, E78.9, G91.29 |
| 119 | V | Synucleinopathy | PD | pos | high | G20.10, G20.90, F02.3, G91.29, Z96.60 |
| 120 | V | Synucleinopathy | PD | pos | high | G20.11, F02.3, R47.1, Z85.4, I44.7, T46.9, G62.9, I95.1, G47.9 |
| 121 | V | Synucleinopathy | PD | pos | intermediate | G20.10, C43.9, G62.9 |
| 122 | V | Synucleinopathy | PD | pos | high | G20.10, F02.3*, G91.29, F06.0, I25.13, I25.15, I10.00, E11.90, E78.9, I95.9 |
| 123 | V | Synucleinopathy | PD | pos | intermediate | G20.10, G23.2 |
| 124 | V | Synucleinopathy | PD | pos | intermediate | G20.10, F02.3, D64.9 |
| 125 | V | Synucleinopathy | PD | neg | intermediate | G20.10, F06.2, F02.3, G47.9, R13.9, R47.1, I95.1, R73.0 |
| 126 | V | Synucleinopathy | PD | pos | intermediate | G20.01, I10.90, R35 |
| 127 | V | Synucleinopathy | MSA-C | pos | high | G23.3, I10.90, N40, R33, F06.7 |
| 128 | V | Synucleinopathy | PD | pos | high | G20.10, F06.0 |
| 129 | V | Synucleinopathy | PD | pos | high | G20.10, G62.9, I11.00, I48.2, M85.89, N40, H91.9, D69.61 |
| 130 | V | Synucleinopathy | PD | pos | high | G20.11, F06.2, F03, F05.9, I10.90, G62.9, M47.99, I25.19, R47.1, R13.9 |
| 131 | V | Synucleinopathy | PD | pos | intermediate | G20.11, F31.3, J44.99, I10.00,F06.7 |
| 132 | V | Synucleinopathy | PD | pos | intermediate | G20.11, F02.3, I10.00, I95.1, F06.0 |
| 133 | V | Synucleinopathy | MSA-P | neg | intermediate | G23.2, L23.5 |
| 134 | V | Synucleinopathy | MSA-P | pos | intemediate | G23.2, G62.9, N40, K59.09, I95.1, R39.1 |
